# The thalamus and its subnuclei: a gateway to obsessive-compulsive disorder

**DOI:** 10.1101/2021.09.06.21262530

**Authors:** findings from the ENIGMA-OCD Working Group, Cees J. Weeland, Selina Kasprzak, Niels T. de Joode, Yoshinari Abe, Pino Alonso, Stephanie H. Ameis, Alan Anticevic, Paul D. Arnold, Srinivas Balachander, Nerisa Banaj, Nuria Bargallo, Marcelo C. Batistuzzo, Francesco Benedetti, Jan C. Beucke, Irene Bollettini, Vilde Brecke, Silvia Brem, Carolina Cappi, Yuqi Cheng, Kang Ik K. Cho, Daniel L. C. Costa, Sara Dallaspezia, Damiaan Denys, Goi Khia Eng, Sónia Ferreira, Jamie D. Feusner, Martine Fontaine, Jean-Paul Fouche, Rachael G. Grazioplene, Patricia Gruner, Mengxin He, Yoshiyuki Hirano, Marcelo Q. Hoexter, Hao Hu, Chaim Huyser, Fern Jaspers-Fayer, Norbert Kathmann, Christian Kaufmann, Minah Kim, Kathrin Koch, Yoo Bin Kwak, Jun Soo Kwon, Luisa Lazaro, Chiang-Shan R. Li, Christine Lochner, Rachel Marsh, Ignacio Martínez-Zalacaín, David Mataix-Cols, Jose M Menchón, Luciano Minnuzi, Pedro Silva Moreira, Pedro Morgado, Akiko Nakagawa, Takashi Nakamae, Janardhanan C. Narayanaswamy, Erika L. Nurmi, Joseph O’Neill, Ana E. Ortiz, Jose C. Pariente, John Piacentini, Maria Picó-Pérez, Federica Piras, Fabrizio Piras, Christopher Pittenger, Y.C. Janardhan Reddy, Daniela Rodriguez-Manrique, Yuki Sakai, Eiji Shimizu, Venkataram Shivakumar, Helen Blair Simpson, Noam Soreni, Carles Soriano-Mas, Nuno Sousa, Gianfranco Spalletta, Emily R. Stern, Michael C. Stevens, S. Evelyn Stewart, Philip R. Szeszko, Jumpei Takahashi, Tais Tanamatis, Anders Lillevik Thorsen, David F. Tolin, Ysbrand D. van der Werf, Hein van Marle, Guido A. van Wingen, Daniela Vecchio, Ganesan Venkatasubramanian, Susanne Walitza, Jicai Wang, Zhen Wang, Anri Watanabe, Lidewij H. Wolters, Xiufeng Xu, Je-Yeon Yun, Qing Zhao, the ENIGMA-OCD working group, Tonya White, Paul .M. Thompson, Dan J. Stein, Odile A. van den Heuvel, Chris Vriend

## Abstract

**Objective:** Higher thalamic volume has been found in children with obsessive-compulsive disorder (OCD) and children with clinical-level symptoms within the general population. Particular thalamic subregions may drive these differences. The ENIGMA-OCD working group conducted mega- and meta-analyses to study thalamic subregional volume in OCD across the lifespan.

**Method:** Structural T_1_-weighted brain magnetic resonance imaging (MRI) scans from 2,649 OCD patients and 2,774 healthy controls across 29 sites (50 datasets) were processed using the FreeSurfer built-in *ThalamicNuclei* pipeline to extract five thalamic subregions. Volume measures were harmonized for site effects using ComBat before running separate multiple linear regression models for children, adolescents, and adults to estimate volumetric group differences. All analyses were preregistered (https://osf.io/73dvy) and adjusted for age, sex and intracranial volume.

**Results:** Unmedicated pediatric OCD patients (< 12 years) had larger lateral (*d* = 0.46), pulvinar (*d* = 0.33), ventral (*d* = 0.35) and whole thalamus (*d* = 0.40) volumes at unadjusted *p*-values <0.05. Adolescent patients showed no volumetric differences. Adult OCD patients compared with controls had smaller volumes across all subregions (anterior, lateral, pulvinar, medial, and ventral) and smaller whole thalamic volume (*d* = -0.15 to -0.07) after multiple comparisons correction, mostly driven by medicated patients and associated with symptom severity. The anterior thalamus was also significantly smaller in patients after adjusting for thalamus size.

**Conclusion:** Our results suggest that OCD-related thalamic volume differences are global and not driven by particular subregions and that the direction of effects are driven by both age and medication status.

## INTRODUCTION

Obsessive-compulsive disorder (OCD) is a debilitating mental disorder with a lifetime prevalence of 1.3% percent that often starts during childhood and follows a chronic course (1). Structural and functional differences in cortico-striato-thalamo-cortical (CSTC) circuits have been linked to commonly observed affective and cognitive deficits, including fear conditioning and extinction, planning ability, response inhibition, habitual behaviour, and emotion regulation (2). The thalamus lies at the core of these functionally segregated CSTC circuits. Previous work from the ENIGMA-OCD Working Group revealed that the thalamus is larger in unmedicated pediatric OCD patients compared with controls (3) and shows more leftward asymmetry (4). The former finding was corroborated in the population-based Generation R study, showing that school-aged children with symptoms above a clinical cut-off had a larger thalamus than symptom-free children (5).

Thalamus structure is not uniform and consists of several nuclei with distinct connectivity and functions (6). The anterior nucleus connects fronto-limbic regions including the medial orbitofrontal and ventromedial prefrontal cortex (7). The mediodorsal nucleus is involved in both emotional and cognitive processing, including reward devaluation, fear extinction, working memory, behavioural flexibility, and goal-directed behaviour (8, 9). The pulvinar nucleus functions as an association nucleus involved in attributing salience to visual stimuli and modulating attention and behaviour accordingly (9, 10). The ventral thalamus includes the ventral anterior and ventral lateral nucleus, which are involved in motor control (9). Functional deficits in all of the aforementioned domains and their associated circuits have been implicated in the neurobiological model of OCD (2). Therefore, higher thalamus volume in OCD may reflect structural or functional changes within these circuits that are expressed in enlarged volume of specific nuclei.

Few studies have investigated OCD-related morphological differences of the thalamus at the subregional level. Shape analysis has found increased surface area of the anterior and pulvinar nuclei in adult OCD patients compared with controls (11). Within school-aged children from the Generation R study, all thalamic nuclei except the pulvinar trended towards larger volumes in the clinical symptom group compared with children without symptoms (12). The ventral nucleus group showed the largest difference, though this was no longer significant after correction for multiple comparisons (12). Another recent study found that adult patients compared with controls had lower volumes of the left posterior thalamus, which includes the pulvinar and geniculate nuclei, and smaller posterior thalamus volume was also associated with a later age of onset of OCD (13). A limitation of previous literature is the focus on a specific age group, limiting the ability to explore differences across the lifespan. The datasets of ENIGMA-OCD allow comparison across a larger age range, including children, adolescents and adults.

We performed mega- and meta-analyses comparing thalamic nuclei volume between OCD patients and healthy controls across 29 sites worldwide. We used the automated segmentation algorithm implemented in FreeSurfer 7.1.1 that uses a histology-based probabilistic atlas created by Iglesias and colleagues (14). We also examined the effects of medication status, age, sex, age of disease onset, and symptom severity. Based on the literature, we expected that pediatric OCD patients would show larger nuclei volumes compared with healthy controls, mostly driven by larger volume of the anterior and ventral thalamus. Furthermore, adult OCD patients were expected to have lower pulvinar volumes compared with healthy controls (13). Based on our previous analyses on subcortical and cortical morphology, we expected that volume enlargement in children would be driven by unmedicated patients and lower volume in adults by medicated patients (3, 15). We also expected that adult OCD patients with a childhood onset would show larger nuclei volumes than patients with adult-onset OCD.

## METHODS

### Samples

The ENIGMA-OCD working group is a collaboration of 29 international research institutes that provided 50 datasets of structural brain imaging and clinical data from OCD patients and control participants without psychopathology. These samples partly overlap with the previous cortical and subcortical mega-analyses performed by this group (3, 15). We divided the participants into three age groups: children under 12 years, adolescents aged 12-17 years, and adults aged 18 years and older. After excluding 45 participants based on poor segmentations, the dataset included 5,423 participants, among which 2,678 OCD patients (96 children, 317 adolescents, 2,265 adults) and 2,746 healthy controls (90 children, 254 adolescents, 2,402 adults). The institutional review boards of each participating site permitted the analysis of measures extracted from coded, de-identified individual participant data.

### Image Acquisition and Processing

T_1_-weighted structural brain magnetic resonance imaging (MRI) images of the participants were acquired at each site and processed using the latest stable release of FreeSurfer (version 7.1.1) for segmentation of the whole thalamus. Next, the pipeline of Iglesias and colleagues (14) was used to segment the thalamus into 25 nuclei, which we grouped into five different subregions per hemisphere: anterior, lateral, ventral, intralaminar/medial and pulvinar (see Figure 1 for an overview of the subdivision). A containerized version (Singularity) of the pipeline was created to facilitate cross-platform compatibility at each site (http://datasets.datalad.org/?dir=/shub/chrisvriend/ENIGMA_subthal) and allow standardized quality inspection of the segmentation by means of custom-made Python and shell scripts. All segmentations were visually inspected and investigated for statistical outliers. Full details of the image exclusion criteria and quality assessments are described in Supplementary Section S1.

**Figure 1:**
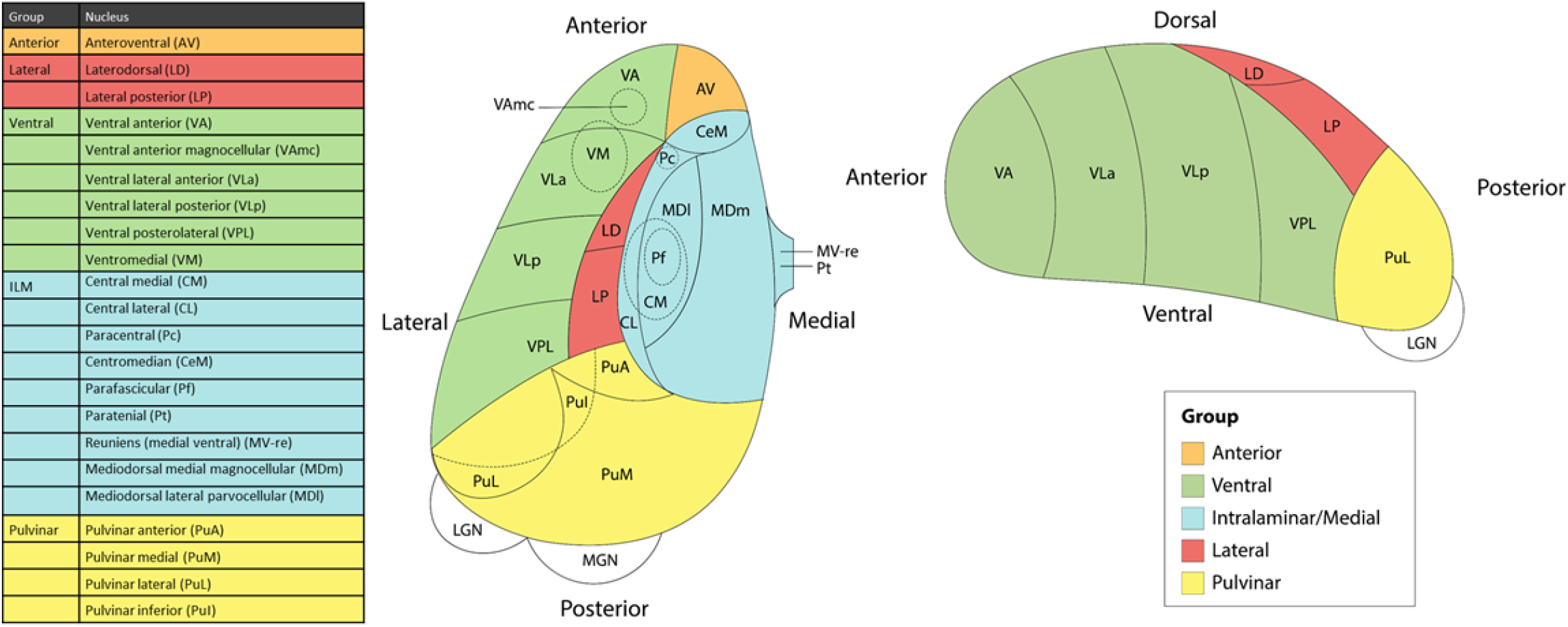
Schematic representation of thalamic nuclei grouping. Note: Figure adapted from *Thalamic Subregions and Obsessive-Compulsive Symptoms in 2,500 Children From the General Population* by Weeland et al., 2021, *Journal of the American Academy of Child and Adolescent Psychiatry*.

### Statistical Analysis

We conducted both mega- and meta-analyses. Statistical analyses were conducted in accordance with statistical protocols of the ENIGMA Consortium (http://enigma.ini.usc.edu/). We pre-registered our analysis plan and hypotheses prior to conducting any analyses (https://osf.io/73dvy). Mega-analyses were performed by pooling the extracted volume measures for each participant across all sites. The data were harmonized to adjust for possible cross-site batch effects using a modified version of the ComBat function (16). Multiple linear regression models were run on the harmonized data. We conducted separate analyses for children, adolescents and adults. The bilateral mean volumes of the five subregions served as the main outcome measures. Diagnosis was used as a binary predictor in all analyses. All models were adjusted for age, sex, and total intracranial volume (ICV). We additionally investigated the interaction of age, age-squared, sex, or sex-by-age. Details of the meta-analysis are described in Supplementary Section S2.

We computed Cohen’s *d* (*d)* effect sizes using the *t*-statistic of the main predictor variable in the regression models. Medication effects were investigated by conducting stratified analyses comparing medicated versus unmedicated OCD patients with the healthy control group. Similarly, we conducted stratified analyses for the adult-onset and child-onset (onset before age 18 years) groups to study the influence of time of onset. We also investigated the continuous association of subregional volume with illness severity, expressed in terms of the total severity score of the adult and child versions of the Yale-Brown Obsessive Compulsive Scale (Y-BOCS) (17, 18). In post-hoc (i.e. not pre-registered) analyses, lateralization of volume differences was assessed in children and adults by computing an asymmetry index ([L – R]/ ([L + R] / 2)) for each subregion (4). Between-group differences between the ratio of each subregion relative to whole thalamus volume were investigated to investigate relative differences in thalamic volume. The main analyses were corrected for multiple comparisons by applying a Benjamini-Hochberg false discovery rate correction (*q* = 0.05) across the five subregions tested. We also report uncorrected results throughout the manuscript.

## RESULTS

Table 1 provides an overview of the demographic and clinical characteristics of the pooled samples for each age group. The demographic and clinical characteristics of the separate samples are displayed in Table S1 in the data supplement.

**Table 1:**
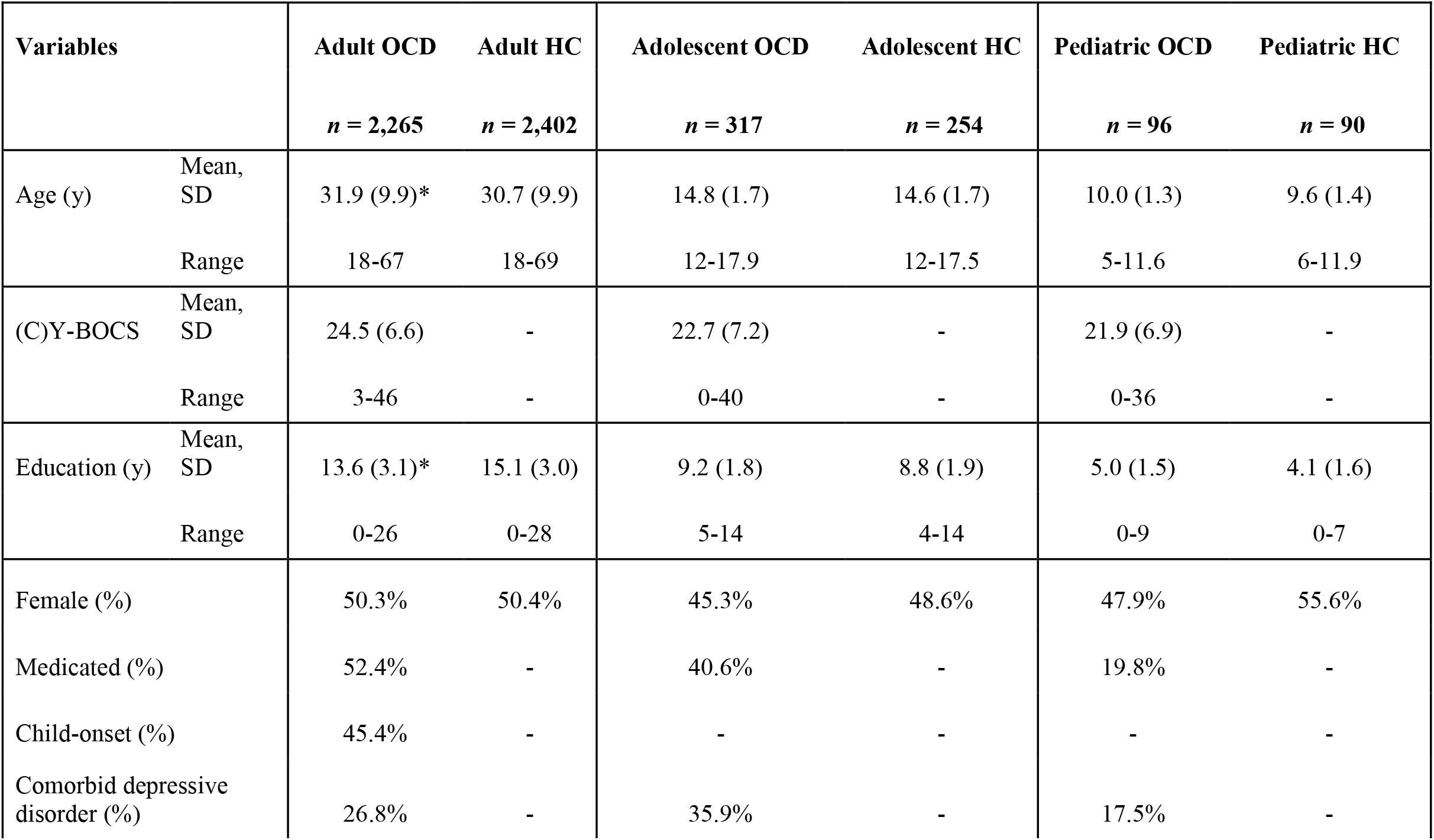
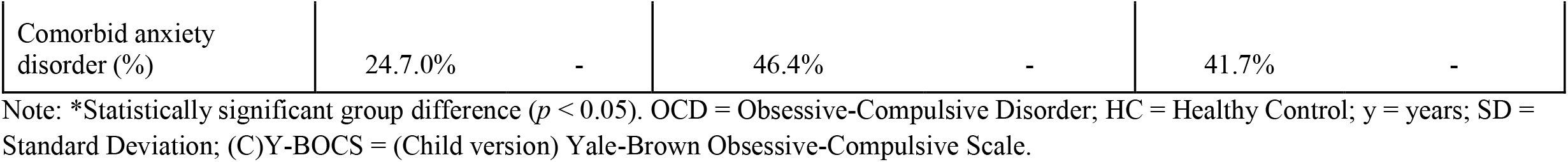
Pooled descriptive statistics of obsessive-compulsive patients and healthy control participants for the adult (age 18 or higher), adolescent (age 12-17) and child (age under 12) age groups.

### Mega-analysis

#### Thalamic subregional volume differences between OCD patients and controls

##### Children (< 12 years)

Consistent with previous findings, whole thalamus volume was larger in OCD patients compared with controls (*d* = 0.32, *p* = 0.03). Larger volume was observed in children with OCD compared with healthy controls in the lateral subregion (*d* = 0.37, *p* = 0.01, *p*_FDR_ = 0.08), but the difference did not survive correction for multiple comparisons. The other subregions trended towards increased volume in OCD patients compared with controls (*d* = 0.16 to 0.28, unadjusted *p* = 0.06 to 0.30) (see Figure 2 and Table 2). We found no significant differences in relative thalamus volume (see Table S2).

**Figure 2:**
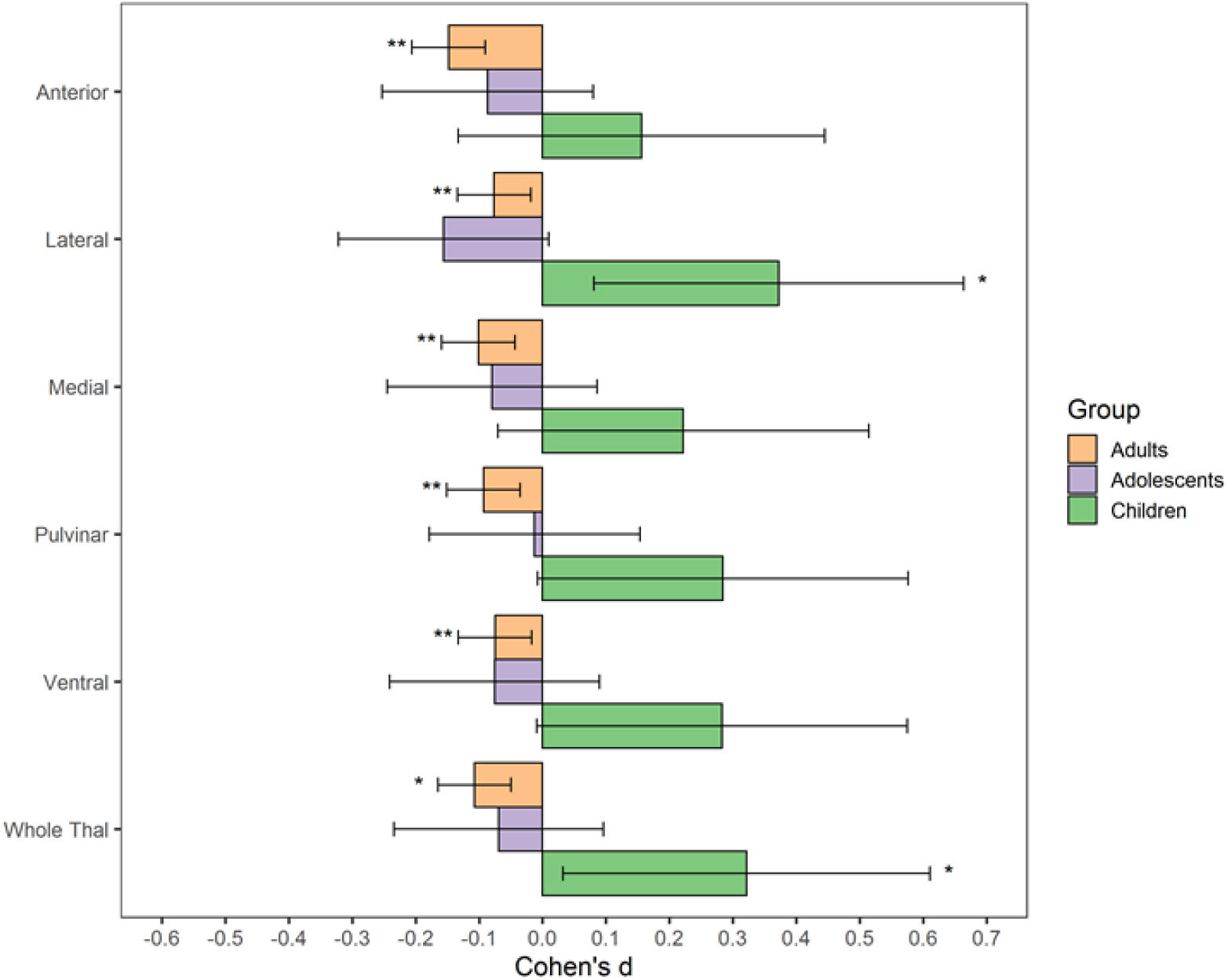
Volumetric differences between obsessive-compulsive patients and healthy controls by age group. Note: \**p*_uncorrected_ < 0.05; * \**p*_FDR_ < 0.05; volumetric differences are adjusted for age, sex and intracranial volume. Thal = Thalamus.

**Table 2:** Multiple linear regression output of volumetric difference between obsessive-compulsive patients and healthy controls for children (< 12 years), adolescents (12-17 years) and adults (age 18 or higher)

|  | Cohen's *d* [95% CI] | *p*-value | *p*-value<br>(FDR) | *n* (OCD) | *n* (HC) |
| --- | --- | --- | --- | --- | --- |
| *Children* |  |  |  |  |  |
| Ventral | 0.28 [-0.009, 0.575] | 0.061 | 0.102 | 94 | 88 |
| Pulvinar | 0.28 [-0.008, 0.576] | 0.060 | 0.102 | 94 | 88 |
| Medial | 0.22 [-0.07, 0.515] | 0.143 | 0.178 | 94 | 87 |
| Lateral | 0.37 [0.081, 0.664] | 0.014 | 0.068 | 95 | 89 |
| Anterior | 0.16 [-0.133, 0.445] | 0.297 | 0.297 | 95 | 90 |
| Whole Thal | 0.32 [0.032, 0.611] | 0.032 | - | 96 | 90 |
| *Adolescents* |  |  |  |  |  |
| Ventral | -0.076 [-0.241, 0.09] | 0.374 | 0.467 | 314 | 253 |
| Pulvinar | -0.012 [-0.178, 0.154] | 0.884 | 0.884 | 312 | 252 |
| Medial | -0.08 [-0.245, 0.086] | 0.348 | 0.467 | 316 | 253 |
| Lateral | -0.16 [-0.322, 0.01] | 0.067 | 0.334 | 316 | 252 |
| Anterior | -0.086 [-0.253, 0.08] | 0.311 | 0.467 | 311 | 251 |
| Whole Thal | -0.069 [-0.234, 0.096] | 0.416 | - | 317 | 254 |

| *Adults* |  |  |  |  |  |
| --- | --- | --- | --- | --- | --- |
| Ventral | -0.075 [-0.133, -0.017] | 0.011 | 0.011 | 2200 | 2383 |
| Pulvinar | -0.093 [-0.151, -0.035] | 0.002 | 0.003 | 2211 | 2381 |
| Medial | -0.10 [-0.159, -0.043] | 0.001 | 0.002 | 2198 | 2390 |
| Lateral | -0.08 [-0.134, -0.019] | 0.010 | 0.011 | 2201 | 2402 |
| Anterior | -0.15 [-0.206, -0.09] | 5.66E-07 | 2.82E-06 | 2195 | 2400 |
| Whole Thal | -0.11 [-0.165, -0.05] | 0.0003 | - | 2265 | 2402 |

Note: Models were adjusted for age, sex, and intracranial volume; 95% CI = 95% Confidence Interval; FDR = False Discovery Rate; n = number of participants; OCD = Obsessive-Compulsive Disorder; HC = Healthy Controls; Thal = Thalamus

##### Adolescents (12-17 years)

There were no significant volume differences of the thalamus as a whole or subregions between adolescent OCD patients and healthy controls (see Figure 2 and Table 2). There was also no group differences in relative thalamus volume (Table S3).

##### Adults (18-69 years)

Whole thalamus volume was smaller in adult OCD patients compared with controls (*d* = -0.11, *p =* 0.001). All subregions had smaller volume in adult OCD patients compared with healthy controls (*d =* -0.07 to -0.15, *p*_*FDR*_ = <0.001 to 0.011, Figure 2 and Table 2) and in the whole thalamus. In addition, anterior volume relative to whole thalamus volume was significantly smaller in adults patients (*d* = -0.11, *p*_*FDR*_ = 0.001, Table S4).

#### Influence of medication status

Figure 3 visualizes the volumetric differences between medicated patients, unmedicated patients and healthy controls in relation to age and sex.

**Figure 3:**
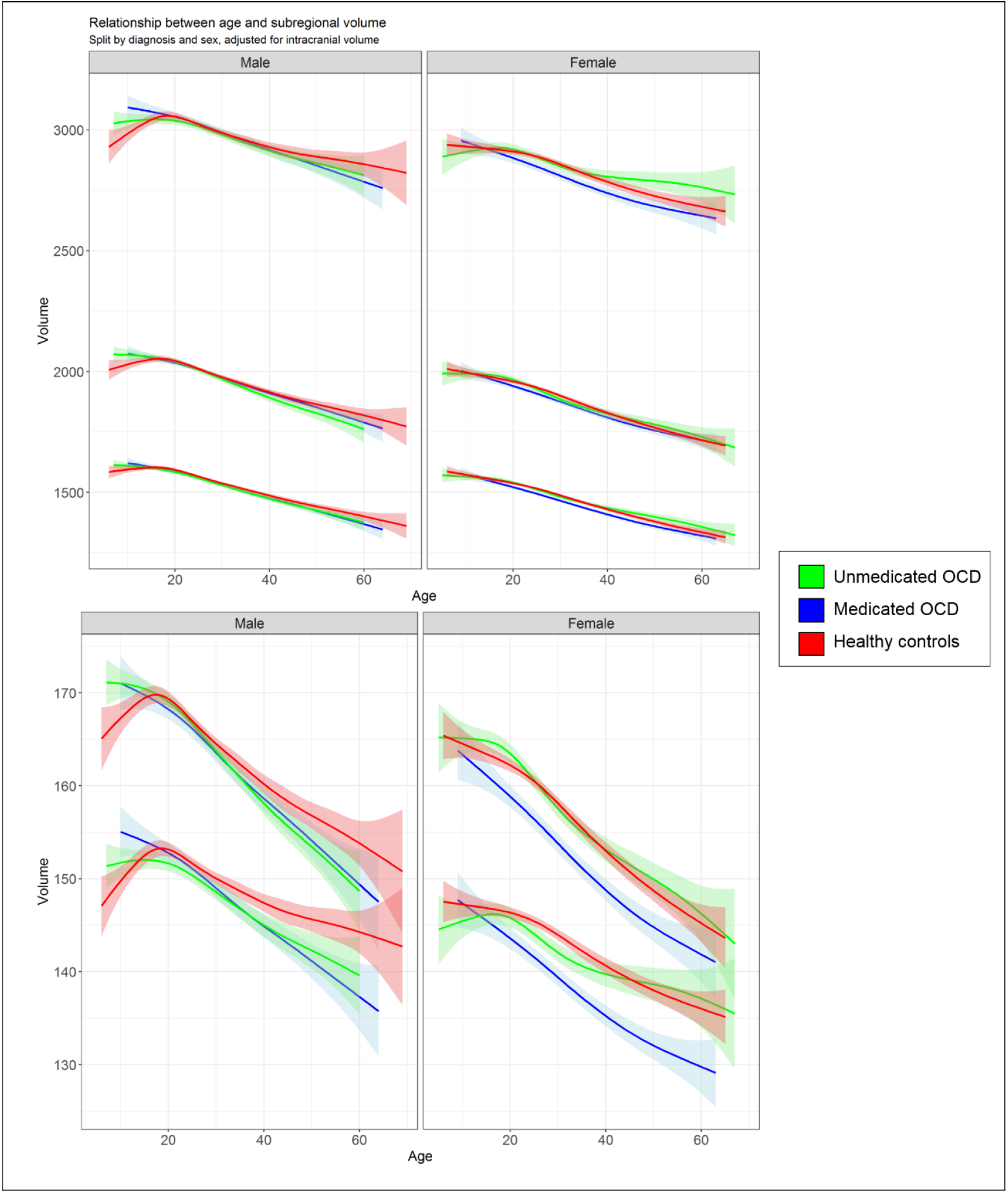
Age trajectory of thalamus subregion volume for medicated obsessive-compulsive patients, unmedicated patients and healthy controls, split by sex and diagnosis. Note: Adjusted for intracranial volume. Shading represents error margins. OCD = Obsessive-Compulsive Disorder.

##### Children

Unmedicated children with OCD compared with healthy controls had a larger lateral volume (*d* = 0.46, *p*_*FDR*_ = 0.02). At an unadjusted significance threshold, patients had a larger ventral subregion (*d* = 0.35, *p* = 0.03) and pulvinar (*d* = 0.33, *p* = 0.03) (see Table S5). We found no significant volume differences between medicated children with OCD and healthy controls or between medicated and unmedicated children with OCD (see Tables S6 and S7).

##### Adolescents

We found no significant differences between medicated adolescents with OCD, unmedicated adolescent OCD patients and healthy controls (see Table S8-10).

##### Adults

Medicated adults with OCD compared with healthy controls exhibited a significantly smaller volume of all thalamic subregions (*d =* -0.09 to -0.17, *p*_*FDR*_ = <0.001 to 0.02) (Table S11), but there were no significant differences between unmedicated adult OCD patients and healthy controls (Table S12) or between medicated and unmedicated patients (Table S13).

#### Influence of age and sex on OCD-related differences in thalamic nuclei volume

Results of the interaction models are displayed in Tables S14-S25. In children, we found an age-by-diagnosis interaction for the medial (Cohen’s *d* = 0.30, *p* = 0.046, *p*_*FDR*_ *=* 0.076), pulvinar (*d* = 0.31, *p* = 0.041, *p*_*FDR*_ *=* 0.076), ventral (*d* = 0.36, *p* = 0.018, *p*_*FDR*_ *=* 0.076) nuclei and for the whole thalamus (*d* = 0.36, *p* = 0.017). This indicates higher volume with age in patients versus lower volume with age in controls (see Table S15). We also found an age-by-sex-by-diagnosis interaction in the anterior (*d* = -0.32, *p*_*FDR*_ *=* 0.045), medial (*d* = -0.40, *p*_*FDR*_ *=* 0.026), pulvinar (*d* = -0.39, *p*_*FDR*_ *=* 0.026), ventral (*d* = -0.34, *p*_*FDR*_ *=* 0.045) subregions and for the whole thalamus (*d* = -0.42, *p* = 0.006) (see Table S17), indicative of higher volume in male pediatric OCD patients with age, but lower volume with age in female OCD patients and healthy controls. We found no such interactions in adolescents (Table S18-S21).

In adults, we found a sex-by-diagnosis interaction in the anterior subregion (*d* = -0.07; *p* = 0.03, *p*_FDR_ = 0.13) (see Table S22), indicating stronger volume differences between OCD patients and healthy controls among females. We found no other interactions (see Tables S23-S25).

#### Influence of time of disease onset on thalamic subregional volume in adults

Compared with healthy controls, adult-onset OCD patients had a smaller medial (*d* = -0.10, *p*_*FDR*_ *=* 0.026) and anterior (*d* = -0.14, *p*_*FDR*_ *=* 0.001) thalamic volume. The ventral and lateral subregions were smaller at unadjusted *p*-value < 0.05 (see Table S26). There were no differences between adult patients with a child-onset and controls (see Table S27) or between child-onset and adult-onset patients (see Table S28).

#### Association between illness severity and thalamic subregional volume

Medication use was associated with higher YBOCS scores (*t* = 2.23, *p* = 0.026) and the presence of comorbid depressive disorders (*t* = 3.13, *p* = 0.002) but not comorbid anxiety disorders (t = 0.59, p = 0.56). We therefore additionally adjusted the severity analyses for medication use and comorbid depressive disorders. There was no relationship between symptom severity and subregional volume in pediatric and adolescent OCD patients (see Table S29 and S30). Conversely, symptom severity in adult OCD patients was negatively associated with volume in all subregions (*r* = -0.04 and -0.06) and whole thalamus volume (*r* = -0.07) (see Table S31).

#### Volume asymmetry group differences between patients and controls

We found no significant thalamic subregional volume asymmetry in child, adolescent and adult OCD patients compared with controls (see Table S32-S34).

### Meta-analysis

Methods and results of the meta-analysis are displayed in Supplementary Section S1 and Table S35 and Figure S1. The results of the meta-analysis corroborate the mega-analysis results.

## DISCUSSION

We performed a mega- and meta-analysis investigating differences in thalamic subregion volume between OCD patients and healthy controls across the lifespan. While unmedicated pediatric patients had *larger* pulvinar, ventral and whole thalamus volumes compared with controls, adult OCD patients had *smaller* volumes across all subregions, which was mostly driven by medicated and adult-onset patients. No volume differences were found in adolescent OCD patients. Our results suggests that OCD-related thalamic volume differences are global and not driven by particular subregions and that the direction of effects are driven by both age and medication status.

Given that specific thalamic subregions play a key role in OCD-relevant circuitry (2), we hypothesized morphometric differences in these subregions in OCD. Thalamic subregions have been more intensively studied in relation to schizophrenia and have repeatedly shown subregional volume differences (19), including recently by using the same Iglesias et al. segmentation tool (20, 21). Participants at high risk of developing schizophrenia with auditory hallucinations had a smaller volume of the medial geniculate nucleus and a steeper volume decrease over time compared with symptom-free controls, accompanied by functional connectivity differences in related circuits (21). This suggests that subregional volume changes might be related to specific symptoms. OCD-related symptom dimensions have also been related to distinct structural changes (22). In the current study we only had data on global symptom severity and not specific symptom dimensions. Given the heterogeneity of the OCD phenotype and the many circuits involved, this could explain why the shrinkage of the thalamus was distributed across all subregions.

Conversely, thalamus volume was larger rather than smaller in the child sample, suggesting another process may be at play. For instance, the larger overall thalamus volume could reflect broad neurodevelopmental differences between patients and controls that predispose to developing OCD. During early childhood and adolescence, thalamo-cortical circuits undergo a process of myelination, synaptic remodelling and pruning that give rise to a mature thalamus (23). Recent work showed that compulsivity and impulsivity is related to reduced myelin-related growth in cortico-striatal regions in adolescents (24). Although disagreement exists on the trajectory of normative thalamic development, relative thalamus size has been described to increase until four years of age (25), followed by either a steady decrease (26) or stabilization (25) during childhood and adolescence. The larger volumes observed in our pediatric OCD patients aged 6-12 years may therefore reflect altered neurodevelopment. The association was strongest in unmedicated children, indicating that medication normalizes thalamic volume. This is consistent with the report by Gilbert et al. (27), that revealed enlargement of the thalamus in pediatric patients that normalized after 12-week treatment with paroxetine. Since neurodevelopment is governed by both genetic (28) and environmental influences (29), longitudinal studies integrating these aspects can offer insight into the altered neurodevelopment in OCD.

Thalamic volume declines with healthy aging (30) and shows enhanced decline in various brain disorders (31, 32). Likewise, OCD patients may exhibit a more rapid decline in thalamic volume through various mechanisms. Glutamatergic neurons fulfil an important role within the CSTC circuits. Several studies have found alterations in glutamatergic systems in OCD, indicating possible glutamatergic dysregulation (33, 34). Excitotoxic effects of glutamate have been proposed as a mechanism for local structural loss in schizophrenia (35). Alternatively, the presence of auto-antibodies in conjunction with increased excitatory metabolites in adult OCD suggest activity of autoimmune processes (34).

Our results suggest that medication status largely drives volumetric differences in adult OCD. Selective serotonin reuptake inhibitors (SSRIs) have been reported to induce both neurogenesis and neuronal elimination in the hippocampus (36, 37). In-vitro studies have linked antidepressant exposure to non-neuronal and neuronal apoptosis or death (38, 39), providing a possible mechanism for volume decline of brain structures. However, to our knowledge such effects have not been described in the thalamus. In another study, a single dose of SSRIs caused widespread decreases in subcortical and cortical connectivity as well as increased thalamic connectivity (40). Perhaps functional alterations induce structural remodelling of the thalamus that result in volumetric changes. However, causal effects cannot be inferred due to the cross-sectional methodology and lack of detailed information on medication status.

Only the anterior subregion was significantly smaller relative to overall thalamic volume in adult OCD patients, suggesting that this difference is superimposed on the overall thalamic difference. The ventral anterior nucleus connects to fronto-limbic regions including the medial orbitofrontal and ventromedial prefrontal cortex (7). Lower relative volume of the anterior nucleus may reflect fronto-limbic disruption, which has been linked to the dysregulated fear responses in OCD, characterized by excessive and poorly controlled fear responses to stimuli (2).

The opposite volumetric effects in children versus adults with OCD suggest a relationship with age. This relationship is visualized in Figure 3 that also demonstrates that age-related differences may be sex-dependent. This is consistent with the significant interactions with age and age-by-sex we found in the pediatric sample. However, we are cautious to interpret the pediatric interaction analyses due to the low sample size and skewed age distribution in this group. Conversely, the sufficiently powered adult sample size did not show a significant interaction with age or age-by-sex. This could indicate that differences in volume trajectories across age between patients and controls that appear in the plot are challenging to model with formal testing. However, we did find a sex-by-diagnosis interaction in adults for the anterior subregion (see Table S3), indicating that OCD-related volume differences are on average stronger in females than males.

As in our prior work, we found the whole thalamus was larger in the pediatric patient group (3). In contrast to our previous work, we split the pediatric group into children (<12 years) and adolescents (12-17 years). We found no group difference between adolescent patients and healthy controls, indicating volume differences are most prominent under the age of 12 years. A novel finding is the smaller whole thalamic volume in adult patients, though a trend towards lower thalamic volume in adult OCD patients was already apparent in our previous analysis (3). This discrepancy is explained by increased power from the larger sample size and use of the ComBat algorithm to adjust for batch effects (16), and the more sensitive thalamic segmentation method (14). Our findings in adults are partly consistent with recent work from Jurng et al. in a sample that was also included in the current analysis (13). While the current study found lower volumes of all subregions in adult OCD patients, Jurng et al. found significant differences only in the posterior thalamus region that includes the pulvinar (13). This difference is mostly likely explained by differences in sample size and the fact that only unmedicated patients were included in the Jurng et al sample.

Strengths of our study include the large sample size, the use of a harmonized and containerized processing protocol and quality inspection procedure, as well as adjustment for batch effects that arise from multi-site data. Furthermore, separation into three age groups allowed us to identify stage-specific volume differences. Nevertheless, the cross-sectional nature of the study complicates identifying developmental processes and medication effects that seem to underlie the observed volume effects. Furthermore, despite adjustment for batch effects, we are not able to overcome all effects that arise from scanner and site differences. Since all sites included both patient and control groups, large effects of site differences are, however, not expected. Although the segmentation atlas used for this analysis was created using an adult sample (14), the atlas was used successfully in previous analyses in pediatric samples (12).

Despite these limitations, our results provide the first robust findings on OCD-related volume differences in the thalamus across different developmental stages. In contrast with our hypotheses, we observed mostly global thalamic rather than region-specific volume differences, suggesting that the relationship with OCD is not driven by changes of a particular subregion but overall thalamus volume. Future endeavours should focus on disentangling longitudinal age-related and medication-related effects to determine what drives these global differences. Furthermore, multimodal approaches including resting-state functional MRI and diffusion-weighted imaging could bolster our understanding of thalamic connectivity and its involvement in OCD, preferably at higher field strengths to attain optimal contrast for high quality images.

## Supporting information

ENIGMA-OCD Working group contributor list - alphabetical

## Data Availability

The data used for the manuscript is not publicly available

## Acknowledgments

The ENIGMA Obsessive-Compulsive Disorder Working Group gratefully acknowledges support from the NIH BD2K award U54 EB020403 (PI: *P*.*M. Thompson*) and Amsterdam Neuroscience (CIA-2019-03-A to *O*.*A. van den Heuvel*, Amsterdam Neuroscience Alliance Project to *G. A. van Wingen*). Supported by the Japan Society for the Promotion of Science (JSPS; KAKENHI Grants No. 18K15523 to *Y. Abe*, No. 19K03308 to *Y. Hirano* and *A. Nakagawa*, No. 16K04342 to *A. Nakagawa*); National Institute of Mental Health (NIMH; Grant no. 5R01MH116038 to *A. Anticevic*, No. R01MH085900 to *J*.*D. Feusner*, No. K23 MH115206 to *P. Gruner*, No. R01MH104648 and No. R21MH101441 to *R. Marsh*, No. R01MH085900 and No. R01MH081864 to *J. O’Neill*, No. 1R01MH081864 to *J. Piacentini*, No. K24 MH121571 to *C. Pittenger*, No. *R01 MH074934*, No. R21/R33MH107589 and No. R01MH111794 to *E*.*R. Stern*, No. R01 MH074934 to *D. Tolin);* the Alberta Innovates Translational Health Chair in Child and Youth Mental Health and the Ontario Brain Institute funding *P*.*D. Arnold*; Swiss National Science Foundation (Grant No. 320030_130237) and the Hartmann Müller Foundation (Grant No. 1460) to *S. Brem*; Fundação Faculdade de Medicina (Grant No. 86.768 to *D*.*L*.*C. Costa*); the Japan Agency for Medical Research and Development (AMED Brain/MINDS Beyond program Grant No. JP21dm0307002 to *Y. Hirano*, No. JP21dm0307008 to *Y. Sakai*); Michael Smith Foundation for Health Research to *F. Jaspers-Fayer*; the Deutsche Forschungsgemeinschaft (DFG; to *D. Rodriguez-Manrique*, Grant No. DFG 815/6-1 to *N. Kathmann*, No. KO 3744/11-1 to *K. Koch*); The Basic Science Research Program through the National Research Foundation of Korea (NRF) (Grant no. 2019R1A2B5B03100844 to *J*.*S. Kwon*); the Marató TV3 Foundation (Grant No. 01/2010 and No. 091710 to *L. Lazaro*); the National Research Foundation of South Africa to *C. Lochner*; the Portuguese Foundation for Science and Technology (Fundação para a Ciência e a Tecnologia; FCT; Project No. UIDB/50026/2020 and No. UIDP/50026/2020 to *P. Morgado* and *N. Sousa*); the ICVS Scientific Microscopy Platform, member of the national infrastructure PPBI - Portuguese Platform of Bioimaging (Grant No. PPBI-POCI-01-0145-FEDER-022122); the Norte Portugal Regional Operational Programme (NORTE 2020; Grant No. NORTE-01-0145-FEDER-000039 to *P. Morgado* and *N. Sousa*), under the PORTUGAL 2020 Partnership Agreement, through the European Regional Development Fund (ERDF); the FLAD Science Award Mental Health 2021 to *P. Morgado and N. Sousa;* the Government of India grants from the Department of Science and Technology (Grants No. SR/S0/HS/0016/2011 to *Y*.*C. Janardhan Reddy*, and DST INSPIRE faculty grant -IFA12-LSBM-26 to *J. C. Narayanaswamy*) and from the Department of Biotechnology (Grants No. BT/PR13334/Med/30/259/2009 to *Y*.*C. Janardhan Reddy*, and No. BT/06/IYBA/2012 to *Dr. Janardhanan C. Narayanaswamy*, and Accelerator Program for Discovery in Brain Disorders using Stem cells (ADBS), Grant No. BT/PR17316/MED/31/326/2015 to *S. Balachander*); the Italian Ministry of Health (Grant No. RC19-20-21 to *Fabrizio Piras*, No. RC18-19-20-21/A to *G. Spalletta*); the Wellcome Trust-DBT India Alliance (Grant No. 500236/Z/11/Z to *G. Venkatasubramanian*, Early Career Fellowship Grant No. IA/CPHE/18/1/503956 *to. V. Shivakumar*); the Carlos III Health Institute (Grant No. PI16/00889 and No. PI19/01171 to *C. Soriano-Mas*); the Departament de Salut, Generalitat de Catalunya (Grant No. PERIS SLT006/17/249 to *C. Soriano-Mas*); the Agència de Gestió d’Ajuts Universitaris i de Recerca (Grant No. 2017 SGR 1262 to *C. Soriano-Mas*); the International OCD Foundation to *P*.*R. Szeszko*; the Helse Vest Health Authority (Grant No. 911754 and No. 911880 to *A*.*L. Thorsen*); the Netherlands Organisation for Health Research and Development (ZonMw; VIDI Grant No. 91717306 to *O*.*A. van den Heuvel*, TOP Grant No. 91211021 to *T. White*); the South African Medical Research Council (SAMRC) to *D*.*J. Stein*; the Dutch Brain Foundation (Nederlandse Hersenstichting; Grant No. HA-2017-00227 to *C. Vriend)*.

