## Supplementary Information for "The thalamus and its subnuclei: a gateway to obsessive-compulsive disorder"

**S1: Segmentation quality assessment**

The lateral and medial geniculate nuclei were excluded from the analyses due to poor segmentation quality. Each site performed detection of statistical outliers (below Q_1_ - 1.5 * IQR or above Q­_3_ + 1.5 * IQR) for each nuclei group. Segmentation quality of these outliers was visually inspected. Secondly, segmentations that deviated too much from the native (aseg) FreeSurfer segmentation of the whole thalamus or showed large overlap with the white matter mask in the aseg file were flagged and visually inspected as well. Poor segmentations were excluded from analyses.

**S2: Meta-analysis**

*Method*

In line with previous work of the ENIGMA-OCD group, we conducted a meta-analysis by pooling the regression statistics from each sample across all sites. Since the pediatric samples within the working group combined both children and adolescent participants, splitting these samples would lead to biased estimates from having too small or unbalanced groups. We therefore only conducted a meta-analysis on the adult samples.

All regression models were computed for each sample separately. We used R statistical package, version 4.0.3 (1), *metafor* package (2) for an inverse variance-weighted random-effect meta-analysis to derive pooled Cohen’s *d* effect size estimates for each nuclei group. All models were fitted using the restricted maximum likelihood method (REML).

*Results*

Adult patients had significantly lower volume in the ventral (*d =* -0.12*, p_FDR_* = 3.49E-04) and medial (*d =* -0.10*, p_FDR_* = 0.017) nucleus (Table S2.1). All other nuclei trended towards lower volumes in patients compared with controls. The whole thalamus was significantly lower in patients compared with controls (*d =* -0.11*, p =* 0.024).

**Table S2.1: Full meta-analytic result of volumetric difference between adult obsessive-compulsive patients and healthy controls**

|  | **Cohen’s *d* [95% CI]** | ***p*-value** | ***p*_FDR_** | **N (OCD)** | **N (HC)** |
| --- | --- | --- | --- | --- | --- |
| Ventral | -0.122 [-0.183, -0.062] | 6.99E-05 | 3.49E-04 | 2200 | 2383 |
| Pulvinar | -0.071 [-0.150, 0.008] | 0.078 | 0.097 | 2211 | 2381 |
| Medial | -0.101 [-0.174, -0.028] | 0.007 | 0.017 | 2198 | 2390 |
| Lateral | -0.085 [-0.171, 0.002] | 0.055 | 0.092 | 2201 | 2402 |
| Anterior | -0.057 [-0.140, 0.026] | 0.180 | 0.180 | 2195 | 2400 |
| Whole Thal | -0.106 [-0.199, -0.014] | 0.024 | - | 2265 | 2402 |

Note: models were adjusted for age, sex, and intracranial volume; 95% CI = 95% Confidence Interval; FDR = False Discovery Rate; Thal = Thalamus.
