## Supplementary Tables & Figures for "The thalamus and its subnuclei: a gateway to obsessive-compulsive disorder"

**Supplementary Section S2: Tables**

| **Table** | **Title** |
| --- | --- |
| S1 | Demographic and clinical characteristics per sample |
| S2 | Multiple linear regression output of relative volume differences between pediatric obsessive-compulsive patients and healthy controls |
| S3 | Multiple linear regression output of relative volume differences between adolescent obsessive-compulsive patients and healthy controls |
| S4 | Multiple linear regression output of relative volume differences between adult obsessive-compulsive patients and healthy controls |
| S5 | Multiple linear regression output of volumetric difference between pediatric unmedicated obsessive-compulsive patients and healthy controls |
| S6 | Multiple linear regression output of volumetric difference between pediatric medicated obsessive-compulsive patients and healthy controls |
| S7 | Multiple linear regression output of volumetric difference between pediatric medicated and unmedicated obsessive-compulsive patients |
| S8 | Multiple linear regression output of volumetric difference between adolescent medicated obsessive-compulsive patients and healthy controls |
| S9 | Multiple linear regression output of volumetric difference between adolescent unmedicated obsessive-compulsive patients and healthy controls |
| S10 | Multiple linear regression output of volumetric difference between adolescent medicated and unmedicated obsessive-compulsive patients |
| S11 | Multiple linear regression output of volumetric difference between adult medicated obsessive-compulsive patients and healthy controls |
| S12 | Multiple linear regression output of volumetric difference between adult unmedicated obsessive-compulsive patients and healthy controls |
| S13 | Multiple linear regression output of volumetric difference between adult medicated and unmedicated obsessive-compulsive patients |
| S14 | Sex-by-diagnosis interaction in pediatric obsessive-compulsive patients and healthy controls |
| S15 | Age-by-diagnosis interaction in pediatric obsessive-compulsive patients and healthy controls |
| S16 | Age-squared-by-diagnosis interaction in pediatric obsessive-compulsive patients and healthy controls |
| S17 | Age-by-sex-by-diagnosis interaction in pediatric obsessive-compulsive patients and healthy controls |
| S18 | Sex-by-diagnosis interaction in adolescent obsessive-compulsive patients and healthy controls |
| S19 | Age-by-diagnosis interaction in adolescent obsessive-compulsive patients and healthy controls |
| S20 | Age-squared-by-diagnosis interaction in adolescent obsessive-compulsive patients and healthy controls |
| S21 | Age-by-sex-by-diagnosis interaction in adolescent obsessive-compulsive patients and healthy controls |
| S22 | Sex-by-diagnosis interaction in adult obsessive-compulsive patients and healthy controls |
| S23 | Age-by-diagnosis interaction in adult obsessive-compulsive patients and healthy controls |
| S24 | Age-squared-by-diagnosis interaction in adult obsessive-compulsive patients and healthy controls |
| S25 | Age-by-sex-by-diagnosis interaction in adult obsessive-compulsive patients and healthy controls |
| S26 | Multiple linear regression output of volumetric difference between adult-onset obsessive-compulsive patients and healthy controls |
| S27 | Multiple linear regression output of volumetric difference between child-onset obsessive-compulsive patients healthy controls |
| S28 | Multiple linear regression output of volumetric difference between adult-onset patients and child-onset obsessive-compulsive patients |
| S29 | Multiple linear regression output of the association between severity (Child version Yale-Brown Obsessive Compulsive Scale) and volume in pediatric obsessive-compulsive patients. |
| S30 | Multiple linear regression output of the association between severity (Child version Yale-Brown Obsessive Compulsive Scale) and volume in adolescent obsessive-compulsive patients. |
| S31 | Multiple linear regression output of the association between severity (Yale-Brown Obsessive Compulsive Scale) and volume in adult obsessive-compulsive patients. |
| S32 | Multiple linear regression output of the asymmetry index differences between pediatric obsessive-compulsive patients and healthy controls |
| S33 | Multiple linear regression output of the asymmetry index differences between adolescent obsessive-compulsive patients and healthy controls |
| S34 | Multiple linear regression output of the asymmetry index differences between adult obsessive-compulsive patients and healthy controls |

**Table S1: Demographic and clinical characteristics per sample**

| PI | City, Country | Set name | Strength | Age (M) | Age (SD) | % Male | Sev (M) | Sev (SD) | %Med | %Dep | %Anx | *n* (total) | *n*(OCD) | *n* (HC) |
| --- | --- | --- | --- | --- | --- | --- | --- | --- | --- | --- | --- | --- | --- | --- |
| Arnold | Ontario, CAN | POND-SickKids | 3T | 12.2 | 2.9 | 40.0 | 22.5 | 4.1 | - | - | - | 50 | 27 | 23 |
| Benedetti | Milan, ITA | Benedetti Milano OSR | 3T | 33.4 | 11.5 | 54.7 | 30.8 | 5.4 | 77.9 | 12.4 | 6.2 | 223 | 113 | 110 |
| Beuke | Berlin, GER I | HUB-1.5T | 1.5T | 32.1 | 9.5 | 49.0 | 19.8 | 7.1 | 38.5 | 37.5 | 28.8 | 208 | 104 | 104 |
|  | Berlin, GER II | HUB-3T | 3T | 32.2 | 9.9 | 43.9 | 22.5 | 5.3 | 45.5 | 66.3 | 22.8 | 230 | 101 | 129 |
| Cheng | Kunming, CHN | - | 1.5T | 28.9 | 8.7 | 49.4 | 26.8 | 6.0 | 100.0 | 18.2 | 43.2 | 83 | 44 | 39 |
| Denys | Amsterdam, NLD I | Reward study | 3T | 36.6 | 10.1 | 33.3 | 26.7 | 5.9 | 59.3 | 25.9 | 7.4 | 51 | 27 | 24 |
|  | Amsterdam, NLD II | SHA-OCD | 3T | 35.4 | 12.6 | 47.5 | 22.1 | 6.2 | 78.9 | 36.8 | 15.8 | 40 | 19 | 21 |
| Gruner | Connecticut, USA | Pediatric OCD | 3T | 14.3 | 2.1 | 54.3 | 26.9 | 4.5 | 52.2 | 39.1 | 43.5 | 46 | 23 | 23 |
| Heuvel van den | Amsterdam, NLD I | VUmc-AGIKO | 1.5T | 32.6 | 8.5 | 34.0 | 22.7 | 6.1 | - | 27.8 | 16.7 | 103 | 54 | 49 |
|  | Amsterdam, NLD II | VUmc-VENI | 3T | 38.4 | 10.7 | 48.1 | 21.5 | 6.1 | - | 19.0 | 33.3 | 79 | 42 | 37 |
| Hirano | Chiba, JPN I | CHB | 3T | 31.3 | 8.8 | 45.0 | 25.6 | 5.9 | 86.8 | 7.9 | 5.3 | 131 | 57 | 74 |
|  | Chiba, JPN II | CHBC | 3T | 13.7 | 2.0 | 53.7 | 25.7 | 6.9 | 52.9 | - | - | 67 | 34 | 33 |
|  | Chiba, JPN III | CHBSRPB | 3T | 30.2 | 12.3 | 48.0 | 24.9 | 5.9 | 92.0 | 8.0 | 12.0 | 102 | 25 | 77 |
| Hoexter | Sao Paulo, BRA I | USP drug naïve 1.5T | 1.5T | 29.6 | 9.1 | 36.4 | 25.4 | 5.2 | 0.0 | 35.0 | 62.5 | 77 | 40 | 37 |
|  | Sao Paulo, BRA II | USP child 3T | 3T | 12.2 | 2.5 | 59.6 | 27.3 | 5.6 | 0.0 | 20.7 | 65.5 | 57 | 29 | 28 |
| Huyser | Amsterdam, NLD | Bascule | 3T | 13.3 | 2.6 | 39.3 | 24.9 | 5.0 | 0.0 | 20.0 | 43.3 | 56 | 30 | 26 |
| Koch | Munich, GER | Munich | 3T | 32.1 | 10.4 | 43.1 | 21.6 | 5.7 | 63.9 | - | - | 232 | 119 | 113 |
| Kvale | Bergen, NOR | Bergen_1 | 3T | 30.4 | 9.3 | 40.3 | 26.5 | 4.3 | 24.4 | 90.2 | 85.4 | 72 | 41 | 31 |
| Kwon | Seoul, KOR I | Seoul I | 1.5T | 25.7 | 6.7 | 62.8 | 26.7 | 6.6 | 12.1 | 47.7 | 53.3 | 196 | 107 | 89 |
|  | Seoul, KOR II | Seoul II | 1.5T | 24.3 | 4.2 | 61.9 | 21.7 | 6.6 | 24.4 | 60.0 | 82.2 | 147 | 45 | 102 |
|  | Seoul, KOR III | Seoul_III | 3T | 26.4 | 6.2 | 64.0 | 23.5 | 6.7 | 97.6 | 39.0 | 43.9 | 86 | 41 | 45 |
| Lazaro | Barcelona, ESP | IDIBAPS | 3T | 14.6 | 2.1 | 56.1 | 19.9 | 7.1 | 70.8 | 4.5 | 23.6 | 164 | 89 | 75 |
| Mataix-Cols | Stockholm, SWE | London | 1.5T | 37.8 | 11.2 | 38.6 | 25.9 | 7.7 | 36.0 | 6.0 | 8.0 | 83 | 50 | 33 |
| Menchón  Soriano Mas | Barcelona, ESP I | IDIBELL15 | 1.5T | 34.9 | 9.6 | 50.2 | 24.6 | 6.4 | 95.0 | 14.4 | 18.1 | 297 | 160 | 137 |
|  | Barcelona, ESP II | IDIBELL3 | 3T | 30.8 | 8.5 | 39.3 | 23.7 | 6.9 | 100.0 | 16.7 | 6.7 | 61 | 30 | 31 |
| Morgado | Braga, POR I | Braga 1.5T | 1.5T | 28.3 | 8.7 | 39.5 | 25.6 | 5.7 | 95.9 | 10.8 | 5.4 | 177 | 74 | 103 |
|  | Braga, POR II | Braga 3T | 3T | 30.0 | 9.6 | 40.6 | 28.6 | 6.9 | 82.6 | 0.0 | 0.0 | 155 | 46 | 109 |
| Nakamae | Kyoto, JPN I | Kyoto 1.5T | 1.5T | 30.7 | 8.6 | 46.3 | 25.0 | 6.5 | 52.3 | 9.1 | 5.7 | 136 | 88 | 48 |
|  | Kyoto, JPN II | Kyoto 3T | 3T | 30.7 | 8.6 | 41.8 | 21.9 | 6.3 | - | - | - | 79 | 37 | 42 |
| Nurmi | California, USA I | AOCD | 3T | 32.4 | 11.3 | 52.7 | 24.7 | 4.3 | 29.2 | 20.8 | 37.5 | 74 | 48 | 26 |
|  | California, USA II | COCD | 3T | 12.9 | 2.7 | 52.1 | 23.9 | 3.9 | 13.8 | 6.9 | 39.7 | 94 | 58 | 36 |
| Pittenger | Connecticut, USA I | Yale 2014 Adult OCD | 3T | 33.7 | 11.3 | 59.4 | 27.3 | 5.8 | 43.8 | 59.4 | 68.8 | 101 | 32 | 69 |
|  | Connecticut, USA II | Yale HCP Prisma | 3T | 30.3 | 9.5 | 52.1 | 22.0 | 5.0 | 34.2 | 18.4 | 18.4 | 71 | 38 | 33 |
|  | Connecticut, USA III | Yale HCP Trio | 3T | 35.5 | 12.7 | 29.8 | 26.8 | 5.2 | - | 43.5 | 34.8 | 47 | 23 | 24 |
| Reddy | Bangalore, IND I | NIMHANS_Adult_1.5T | 1.5T | 26.7 | 6.5 | 68.0 | 25.8 | 8.2 | - | 22.0 | 16.0 | 97 | 50 | 47 |
|  | Bangalore, IND II | NIMHANS Adult 3T | 3T | 28.2 | 6.2 | 57.8 | 26.0 | 6.3 | 36.3 | 30.3 | 8.1 | 436 | 234 | 202 |
|  | Bangalore, IND III | NIMHANS Child 3T | 3T | 13.7 | 2.1 | 53.3 | 22.4 | 7.1 | 86.7 | 6.7 | 26.7 | 30 | 15 | 15 |
| Simpson | New York, USA I | OCD Circuits R01 | 3T | 28.8 | 7.4 | 51.9 | 24.7 | 4.4 | - | 2.3 | - | 79 | 43 | 36 |
|  | New York, USA II | OCD R21 | 3T | 28.5 | 7.9 | 51.5 | 25.4 | 3.5 | 100.0 | 100.0 | 100.0 | 66 | 33 | 33 |
|  | New York, USA III | Pediatric OCD | 3T | 11.7 | 3.3 | 47.3 | 24.3 | 5.1 | - | - | 46.4 | 55 | 28 | 27 |
| Soreni | Ontario, CAN | OCD MACMASTER | 3T | 12.1 | 2.1 | 51.6 | 21.4 | 7.1 | - | - | 16.7 | 31 | 18 | 13 |
| Spalletta | Rome, ITA I | FSL Rome 1 | 3T | 36.8 | 11.1 | 62.6 | 23.3 | 8.9 | 85.4 | 11.0 | 1.2 | 195 | 82 | 113 |
|  | Rome, ITA II | FSL Rome 2 | 3T | 34.3 | 10.6 | 55.4 | 24.8 | 7.8 | - | - | 11.8 | 56 | 17 | 39 |
| Stein | Cape Town, ZAF | Cape_Town | 3T | 31.4 | 10.4 | 41.3 | 23.1 | 4.2 | 40.9 | - | 4.5 | 46 | 22 | 24 |
| Stern | New York, USA I | SEQ1NKI | 3T | 33.5 | 12.6 | 37.6 | 23.8 | 5.5 | 50.6 | 5.2 | 50.6 | 93 | 77 | 16 |
|  | New York, USA II | SEQ2SEN | 3T | 27.4 | 6.6 | 33.3 | 24.4 | 5.5 | 50.0 | 6.3 | 56.3 | 27 | 16 | 11 |
|  | New York, USA III | SEQ2UFA | 3T | 31.6 | 10.6 | 49.2 | 22.5 | 9.7 | 50.0 | - | 16.7 | 61 | 6 | 55 |
| Stewart | British Columbia, CAN | SESOCD | 3T | 13.9 | 3.1 | 40.4 | 13.2 | 6.7 | 77.8 | 3.7 | 33.3 | 57 | 27 | 30 |
| Tolin | Connecticut, USA | IOLOCD | 3T | 41.5 | 14.7 | 40.0 | 22.6 | 4.8 | 56.0 | 32.0 | 32.0 | 55 | 25 | 30 |
| Walitza | Zurich, CHE I | UZH | 3T | 23.5 | 10.4 | 52.9 | 15.9 | 10.1 | 57.6 | 9.1 | 33.3 | 70 | 33 | 37 |
| Wang | Shanghai, CHN | Zhen Wang OCD | 3T | 27.5 | 8.9 | 55.8 | 25.3 | 4.9 | 1.8 | - | - | 95 | 57 | 38 |

Note: Anx = comorbid anxiety symptoms; Dep = comorbid depression symptoms; HC = healthy controls; OCD = obsessive-compulsive disorder; PI = principal investigator; Sev = severity measured by (Child version)Yale-Brown Obsessive Compulsive Scale; M = mean; SD = standard deviation.

**Table S2: Multiple linear regression output of relative volume differences between pediatric obsessive-compulsive patients and healthy controls**

|  | **Cohen's *d* [95% CI]** | ***p*-value** | ***p*_FDR_** | **n (OCD)** | **n (HC)** |
| --- | --- | --- | --- | --- | --- |
| Ventral | -0.002 [-0.293, 0.288] | 0.987 | 0.987 | 94 | 88 |
| Pulvinar | 0.011 [-0.280, 0.302] | 0.941 | 0.987 | 94 | 88 |
| Medial | -0.115 [-0.407, 0.177] | 0.446 | 0.987 | 94 | 87 |
| Lateral | 0.275 [-0.015, 0.566] | 0.067 | 0.337 | 95 | 89 |
| Anterior | -0.052 [-0.341, 0.236] | 0.727 | 0.987 | 95 | 90 |

Note: models were adjusted for age, sex, and intracranial volume; 95% CI = 95% Confidence Interval; FDR = False Discovery Rate; n = number of participants; OCD = Obsessive-Compulsive Disorder; HC = Healthy Controls.

**Table S3: Multiple linear regression output of relative volume differences between adolescent obsessive-compulsive patients and healthy controls**

|  | **Cohen's *d* [95% CI]** | ***p*-value** | ***p*_FDR_** | **n (OCD)** | **n (HC)** |
| --- | --- | --- | --- | --- | --- |
| Ventral | -0.044 [-0.210, 0.121] | 0.603 | 0.754 | 314 | 253 |
| Pulvinar | 0.074 [-0.092, 0.240] | 0.388 | 0.754 | 312 | 252 |
| Medial | -0.004 [-0.170, 0.161] | 0.959 | 0.959 | 316 | 253 |
| Lateral | -0.146 [-0.312, 0.019] | 0.085 | 0.425 | 316 | 252 |
| Anterior | -0.062 [-0.229, 0.104] | 0.464 | 0.754 | 311 | 251 |

Note: models were adjusted for age, sex, and intracranial volume; 95% CI = 95% Confidence Interval; FDR = False Discovery Rate; n = number of participants; OCD = Obsessive-Compulsive Disorder; HC = Healthy Controls.

**Table S4: Multiple linear regression output of relative volume differences between adult obsessive-compulsive patients and healthy controls**

|  | **Cohen's *d* [95% CI]** | ***p*-value** | ***p*_FDR_** | **n (OCD)** | **n (HC)** |
| --- | --- | --- | --- | --- | --- |
| Ventral | 0.044 [-0.014, 0.102] | 0.141 | 0.353 | 2200 | 2382 |
| Pulvinar | 0.000 [-0.057, 0.058] | 0.988 | 0.988 | 2211 | 2380 |
| Medial | 0.003 [-0.055, 0.061] | 0.927 | 0.988 | 2198 | 2389 |
| Lateral | -0.031 [-0.089, 0.027] | 0.292 | 0.486 | 2201 | 2403 |
| Anterior | -0.108 [-0.166, -0.050] | 2.530E-04 | 0.001 | 2195 | 2399 |

Note: models were adjusted for age, sex, and intracranial volume; 95% CI = 95% Confidence Interval; FDR = False Discovery Rate; n = number of participants; OCD = Obsessive-Compulsive Disorder; HC = Healthy Controls.

**Table S5: Multiple linear regression output of volumetric difference between pediatric unmedicated obsessive-compulsive patients and healthy controls**

|  | **Cohen's *d* [95% CI]** | ***p*-value** | ***p*_FDR_** | **n (OCD)** | **n (HC)** |
| --- | --- | --- | --- | --- | --- |
| Ventral | 0.350 [0.041, 0.662] | 0.029 | 0.067 | 75 | 88 |
| Pulvinar | 0.330 [0.020, 0.64] | 0.040 | 0.067 | 75 | 88 |
| Medial | 0.300 [-0.009, 0.612] | 0.061 | 0.077 | 75 | 87 |
| Lateral | 0.46 [0.145, 0.765] | 0.005 | 0.023 | 76 | 89 |
| Anterior | 0.24 [-0.066, 0.547] | 0.130 | 0.130 | 76 | 90 |
| Whole Thal | 0.400 [0.096, 0.71] | 0.011 | - | 77 | 90 |

Note: models were adjusted for age, sex, and intracranial volume; 95% CI = 95% Confidence Interval; FDR = False Discovery Rate; *n* = number of participants; OCD = Obsessive-Compulsive Disorder; HC = Healthy Controls; Thal = Thalamus.

**Table S6: Multiple linear regression output of volumetric difference between pediatric medicated obsessive-compulsive patients and healthy controls**

|  | **Cohen's *d* [95% CI]** | ***p*-value** | ***p*_FDR_** | ***n* (OCD)** | ***n* (HC)** |
| --- | --- | --- | --- | --- | --- |
| Ventral | 0.22 [-0.276, 0.717] | 0.396 | 0.834 | 19 | 88 |
| Pulvinar | 0.28 [-0.218, 0.777] | 0.283 | 0.834 | 19 | 88 |
| Medial | 0.071 [-0.425, 0.567] | 0.785 | 0.834 | 19 | 87 |
| Lateral | 0.140 [-0.352, 0.639] | 0.581 | 0.834 | 19 | 89 |
| Anterior | -0.054 [-0.549, 0.441] | 0.834 | 0.834 | 19 | 90 |
| Whole Thal | 0.22 [-0.277, 0.715] | 0.399 | - | 19 | 90 |

Note: models were adjusted for age, sex, and intracranial volume; 95% CI = 95% Confidence Interval; FDR = False Discovery Rate; *n* = number of participants; OCD = Obsessive-Compulsive Disorder; HC = Healthy Controls; Thal = Thalamus.

**Table S7: Multiple linear regression output of volumetric difference between pediatric medicated and unmedicated obsessive-compulsive patients**

|  | **Cohen's *d* [95% CI]** | ***p*-value** | ***p*_FDR_** | ***n* (Med)** | ***n* (Unmed)** |
| --- | --- | --- | --- | --- | --- |
| Ventral | -0.37 [-0.88, 0.132] | 0.160 | 0.226 | 19 | 75 |
| Pulvinar | -0.23 [-0.736, 0.273] | 0.382 | 0.382 | 19 | 75 |
| Medial | -0.45 [-0.959, 0.057] | 0.091 | 0.226 | 19 | 75 |
| Lateral | -0.36 [-0.861, 0.15] | 0.181 | 0.226 | 19 | 76 |
| Anterior | -0.42 [-0.926, 0.086] | 0.115 | 0.226 | 19 | 76 |
| Whole Thal | -0.42 [-0.923, 0.088] | 0.116 | - | 19 | 77 |

Note: models were adjusted for age, sex, and intracranial volume; 95% CI = 95% Confidence Interval; FDR = False Discovery Rate; *n* = number of participants; Med = Medicated OCD; Unmed = Unmedicated OCD; OCD = Obsessive-Compulsive Disorder; Thal = Thalamus.

**Table S8: Multiple linear regression output of volumetric difference between adolescent medicated obsessive-compulsive patients and healthy controls**

|  | **Cohen's *d* [95% CI]** | ***p*-value** | ***p*_FDR_** | ***n* (OCD)** | ***n* (HC)** |
| --- | --- | --- | --- | --- | --- |
| Ventral | -0.078 [-0.267, 0.112] | 0.426 | 0.710 | 185 | 253 |
| Pulvinar | 0.036 [-0.154, 0.226] | 0.710 | 0.710 | 184 | 252 |
| Medial | -0.039 [-0.228, 0.151] | 0.692 | 0.710 | 186 | 253 |
| Lateral | -0.096 [-0.285, 0.094] | 0.327 | 0.710 | 185 | 252 |
| Anterior | -0.150 [-0.340, 0.041] | 0.127 | 0.636 | 184 | 251 |
| Whole Thal | -0.051 [-0.24, 0.138] | 0.601 | - | 186 | 254 |

Note: models were adjusted for age, sex, and intracranial volume; 95% CI = 95% Confidence Interval; FDR = False Discovery Rate; *n* = number of participants; OCD = Obsessive-Compulsive Disorder; HC = Healthy Controls; Thal = Thalamus.

**Table S9: Multiple linear regression output of volumetric difference between adolescent unmedicated obsessive-compulsive patients and healthy controls**

|  | **Cohen's *d* [95% CI]** | ***p*-value** | ***p*_FDR_** | ***n* (OCD)** | ***n* (HC)** |
| --- | --- | --- | --- | --- | --- |
| Ventral | -0.099 [-0.313, 0.115] | 0.368 | 0.494 | 126 | 253 |
| Pulvinar | -0.094 [-0.308, 0.121] | 0.395 | 0.494 | 125 | 252 |
| Medial | -0.150 [-0.366, 0.061] | 0.166 | 0.414 | 127 | 253 |
| Lateral | -0.270 [-0.487, -0.059] | 0.013 | 0.064 | 128 | 252 |
| Anterior | -0.036 [-0.251, 0.178] | 0.743 | 0.743 | 125 | 251 |
| Whole Thal | -0.120 [-0.333, 0.092] | 0.271 | - | 128 | 254 |

Note: models were adjusted for age, sex, and intracranial volume; 95% CI = 95% Confidence Interval; FDR = False Discovery Rate; *n* = number of participants; OCD = Obsessive-Compulsive Disorder; HC = Healthy Controls; Thal = Thalamus.

**Table S10: Multiple linear regression output of volumetric difference between adolescent medicated and unmedicated obsessive-compulsive patients**

|  | **Cohen's *d* [95% CI]** | ***p*-value** | | ***p*_FDR_** | ***n* (Med)** | | | ***n* (Unmed)** |
| --- | --- | --- | --- | --- | --- | --- | --- | --- |
| Ventral | -0.028 [-0.255, 0.198] | 0.809 | | 0.809 | 126 | | | 185 |
| Pulvinar | -0.150 [-0.377, 0.078] | 0.202 | | 0.308 | 125 | | | 184 |
| Medial | -0.150 [-0.378, 0.074] | 0.191 | | 0.308 | 127 | | | 186 |
| Lateral | -0.180 [-0.406, 0.046] | 0.121 | | 0.308 | 128 | | | 185 |
| Anterior | 0.140 [-0.092, 0.363] | 0.246 | | 0.308 | 125 | | | 184 |
| Whole Thal | -0.090 [-0.315, 0.135] | 0.437 | - | | | 128 | 186 | |

Note: models were adjusted for age, sex, and intracranial volume; 95% CI = 95% Confidence Interval; FDR = False Discovery Rate; *n* = number of participants; Med = Medicated OCD; Unmed = Unmedicated OCD; OCD = Obsessive-Compulsive Disorder; Thal = Thalamus

**Table S11: Multiple linear regression output of volumetric difference between adult medicated obsessive-compulsive patients and healthy controls**

|  | **Cohen's *d* [95% CI]** | ***p*-value** | ***p*_FDR_** | ***n* (OCD)** | ***n* (HC)** |
| --- | --- | --- | --- | --- | --- |
| Ventral | -0.094 [-0.166, -0.022] | 0.010 | 0.013 | 1088 | 2383 |
| Pulvinar | -0.086 [-0.157, -0.014] | 0.019 | 0.019 | 1097 | 2381 |
| Medial | -0.120 [-0.186, -0.043] | 0.002 | 0.004 | 1094 | 2390 |
| Lateral | -0.094 [-0.166, -0.022] | 0.010 | 0.013 | 1090 | 2402 |
| Anterior | -0.170 [-0.238, -0.095] | 5.46E-06 | 2.73E-05 | 1091 | 2400 |
| Whole Thal | -0.120 [-0.186, -0.044] | 0.001 | - | 1110 | 2402 |

Note: models were adjusted for age, sex, and intracranial volume; 95% CI = 95% Confidence Interval; FDR = False Discovery Rate; *n* = number of participants; OCD = Obsessive-Compulsive Disorder; HC = Healthy Controls; Thal = Thalamus.

**Table S12: Multiple linear regression output of volumetric difference between adult unmedicated obsessive-compulsive patients and healthy controls**

|  | **Cohen's *d* [95% CI]** | ***p*-value** | ***p*_FDR_** | ***n*N (OCD)** | ***n* (HC)** |
| --- | --- | --- | --- | --- | --- |
| Ventral | -0.026 [-0.100, 0.048] | 0.493 | 0.493 | 987 | 2383 |
| Pulvinar | -0.085 [-0.159, -0.010] | 0.026 | 0.064 | 988 | 2381 |
| Medial | -0.063 [-0.137, 0.011] | 0.098 | 0.163 | 978 | 2390 |
| Lateral | -0.036 [-0.110, 0.038] | 0.345 | 0.431 | 988 | 2402 |
| Anterior | -0.092 [-0.166, -0.018] | 0.015 | 0.064 | 981 | 2400 |
| Whole Thal | -0.071 [-0.145, 0.003] | 0.059 | - | 1000 | 2402 |

Note: models were adjusted for age, sex, and intracranial volume; 95% CI = 95% Confidence Interval; FDR = False Discovery Rate; n = number of participants; OCD = Obsessive-Compulsive Disorder; HC = Healthy Controls; Thal = Thalamus.

**Table S13: Multiple linear regression output of volumetric difference between adult medicated and unmedicated obsessive-compulsive patients**

|  | **Cohen's *d* [95% CI]** | ***p*-value** | ***p*_FDR_** | ***n* (Med)** | ***n* (Unmed)** |
| --- | --- | --- | --- | --- | --- |
| Ventral | -0.070 [-0.156, 0.016] | 0.112 | 0.280 | 1088 | 987 |
| Pulvinar | -0.005 [-0.091, 0.081] | 0.907 | 0.907 | 1097 | 988 |
| Medial | -0.052 [-0.138, 0.035] | 0.241 | 0.302 | 1094 | 978 |
| Lateral | -0.055 [-0.141, 0.031] | 0.208 | 0.302 | 1090 | 988 |
| Anterior | -0.071 [-0.157, 0.016] | 0.109 | 0.280 | 1091 | 981 |
| Whole Thal | -0.046 [-0.132, 0.039] | 0.287 | - | 1110 | 1000 |

Note: models were adjusted for age, sex, and intracranial volume; 95% CI = 95% Confidence Interval; FDR = False Discovery Rate; *n* = number of participants; Med = Medicated OCD; Unmed = Unmedicated OCD; OCD = Obsessive-Compulsive Disorder; Thal = Thalamus.

**Table S14: Sex-by-diagnosis interaction in pediatric obsessive-compulsive patients and healthy controls**

|  | **Cohen’s *d* [95% CI]** | ***p*-value** | ***p*_FDR_** | ***n* (OCD)** | ***n* (HC)** |
| --- | --- | --- | --- | --- | --- |
| Ventral | -0.120 [-0.410, 0.172] | 0.431 | 0.718 | 94 | 88 |
| Pulvinar | -0.160 [-0.454, 0.129] | 0.283 | 0.718 | 94 | 88 |
| Medial | -0.140 [-0.433, 0.151] | 0.354 | 0.718 | 94 | 87 |
| Lateral | -0.070 [-0.359, 0.219] | 0.64 | 0.800 | 95 | 89 |
| Anterior | -0.033 [-0.322, 0.255] | 0.825 | 0.825 | 95 | 90 |
| Whole Thal | -0.130 [-0.420, 0.155] | 0.375 | - | 96 | 90 |

Note: models were adjusted for age, sex, and intracranial volume; 95% CI = 95% Confidence Interval; FDR = False Discovery Rate; *n* = number of participants; OCD = Obsessive-Compulsive Disorder; HC = Healthy Controls; Thal = Thalamus.

**Table S15: Age-by-diagnosis interaction in pediatric obsessive-compulsive patients and healthy controls**

|  | **Cohen’s *d* [95% CI]** | ***p*-value** | ***p*_FDR_** | ***n* (OCD)** | ***n* (HC)** |
| --- | --- | --- | --- | --- | --- |
| Ventral | 0.360 [0.068, 0.654] | 0.018 | 0.076 | 94 | 88 |
| Pulvinar | 0.310 [0.018, 0.603] | 0.041 | 0.076 | 94 | 88 |
| Medial | 0.300 [0.011, 0.598] | 0.046 | 0.076 | 94 | 87 |
| Lateral | 0.058 [-0.231, 0.348] | 0.697 | 0.697 | 95 | 89 |
| Anterior | 0.180 [-0.104, 0.473] | 0.219 | 0.273 | 95 | 90 |
| Whole Thal | 0.360 [0.068, 0.648] | 0.017 | - | 96 | 90 |

Note: models were adjusted for age, sex, and intracranial volume; 95% CI = 95% Confidence Interval; FDR = False Discovery Rate; *n* = number of participants; OCD = Obsessive-Compulsive Disorder; HC = Healthy Controls; Thal = Thalamus

**Table S16: Age-squared-by-diagnosis interaction in pediatric obsessive-compulsive patients and healthy controls**

|  | | **Cohen’s *d* [95% CI]** | ***p*-value** | ***p*_FDR_** | ***n* (OCD)** | ***n* (HC)** |
| --- | --- | --- | --- | --- | --- | --- |
| Ventral | 0.36 [0.068, 0.654] | | 0.018 | 0.076 | 94 | 88 |
| Pulvinar | 0.31 [0.018, 0.603] | | 0.041 | 0.076 | 94 | 88 |
| Medial | 0.3 [0.011, 0.598] | | 0.046 | 0.076 | 94 | 87 |
| Lateral | 0.058 [-0.231, 0.348] | | 0.697 | 0.697 | 95 | 89 |
| Anterior | 0.18 [-0.104, 0.473] | | 0.219 | 0.273 | 95 | 90 |
| Whole Thal | 0.36 [0.068, 0.648] | | 0.017 | - | 96 | 90 |

Note: models were adjusted for age, sex, and intracranial volume; 95% CI = 95% Confidence Interval; FDR = False Discovery Rate; *n* = number of participants; OCD = Obsessive-Compulsive Disorder; HC = Healthy Controls; Thal = Thalamus.

**Table S17 : Age-by-sex-by-diagnosis interaction in pediatric obsessive-compulsive patients and healthy controls**

|  | **Cohen’s *d* [95% CI]** | ***p*-value** | ***p*_FDR_** | ***n* (OCD)** | ***n* (HC)** |
| --- | --- | --- | --- | --- | --- |
| Ventral | -0.340 [-0.632, -0.046] | 0.027 | 0.045 | 94 | 88 |
| Pulvinar | -0.390 [-0.687, -0.100] | 0.010 | 0.026 | 94 | 88 |
| Medial | -0.400 [-0.699, -0.109] | 0.009 | 0.026 | 94 | 87 |
| Lateral | -0.230 [-0.519, 0.061] | 0.132 | 0.132 | 95 | 89 |
| Anterior | -0.320 [-0.609, -0.029] | 0.036 | 0.045 | 95 | 90 |
| Whole Thal | -0.420 [-0.712, -0.131] | 0.006 | - | 96 | 90 |

Note: models were adjusted for age, sex, and intracranial volume; 95% CI = 95% Confidence Interval; FDR = False Discovery Rate; *n* = number of participants; OCD = Obsessive-Compulsive Disorder; HC = Healthy Controls; Thal = Thalamus.

**Table S18: Sex-by-diagnosis interaction in adolescent obsessive-compulsive patients and healthy controls**

|  | **Cohen’s *d* [95% CI]** | ***p*-value** | ***p*_FDR_** | ***n* (OCD)** | ***n* (HC)** |
| --- | --- | --- | --- | --- | --- |
| Ventral | -0.088 [-0.254, 0.077] | 0.298 | 0.703 | 314 | 253 |
| Pulvinar | -0.013 [-0.179, 0.153] | 0.874 | 0.874 | 312 | 252 |
| Medial | -0.068 [-0.234, 0.097] | 0.422 | 0.703 | 316 | 253 |
| Lateral | -0.130 [-0.292, 0.040] | 0.139 | 0.694 | 316 | 252 |
| Anterior | -0.049 [-0.215, 0.118] | 0.568 | 0.710 | 311 | 251 |
| Whole Thal | -0.072 [-0.237, 0.094] | 0.398 | - | 317 | 254 |

Note: models were adjusted for age, sex, and intracranial volume; 95% CI = 95% Confidence Interval; FDR = False Discovery Rate; *n* = number of participants; OCD = Obsessive-Compulsive Disorder; HC = Healthy Controls; Thal = Thalamus

**Table S19: Age-by-diagnosis interaction in adolescent obsessive-compulsive patients and healthy controls**

|  | **Cohen’s *d* [95% CI]** | | ***p*-value** | **p_FDR_** | ***n* (OCD)** | ***n* (HC)** |
| --- | --- | --- | --- | --- | --- | --- |
| Ventral | | 0.025 [-0.141, 0.190] | 0.772 | 0.906 | 314 | 253 |
| Pulvinar | | 0.059 [-0.107, 0.225] | 0.487 | 0.906 | 312 | 252 |
| Medial | | 0.100 [-0.062, 0.269] | 0.222 | 0.906 | 316 | 253 |
| Lateral | | 0.010 [-0.155, 0.176] | 0.906 | 0.906 | 316 | 252 |
| Anterior | | -0.043 [-0.209, 0.123] | 0.614 | 0.906 | 311 | 251 |
| Whole Thal | | 0.067 [-0.098, 0.233] | 0.426 | - | 317 | 254 |

Note: models were adjusted for age, sex, and intracranial volume; 95% CI = 95% Confidence Interval; FDR = False Discovery Rate; *n* = number of participants; OCD = Obsessive-Compulsive Disorder; HC = Healthy Controls; Thal = Thalamus

**Table S20: Age-squared-by-diagnosis interaction in adolescent obsessive-compulsive patients and healthy controls**

|  | **Cohen’s *d* [95% CI]** | ***p*-value** | ***p*_FDR_** | ***n* (OCD)** | ***n* (HC)** |
| --- | --- | --- | --- | --- | --- |
| Ventral | 0.025 [-0.141, 0.190] | 0.772 | 0.906 | 314 | 253 |
| Pulvinar | 0.059 [-0.107, 0.225] | 0.487 | 0.906 | 312 | 252 |
| Medial | 0.100 [-0.062, 0.269] | 0.222 | 0.906 | 316 | 253 |
| Lateral | 0.010 [-0.155, 0.176] | 0.906 | 0.906 | 316 | 252 |
| Anterior | -0.043 [-0.209, 0.123] | 0.614 | 0.906 | 311 | 251 |
| Whole Thal | 0.067 [-0.098, 0.233] | 0.426 | - | 317 | 254 |

Note: models were adjusted for age, sex, and intracranial volume; 95% CI = 95% Confidence Interval; FDR = False Discovery Rate; *n* = number of participants; OCD = Obsessive-Compulsive Disorder; HC = Healthy Controls; Thal = Thalamus

**Table S21: Age-by-sex-by-diagnosis interaction in adolescent obsessive-compulsive patients and healthy controls**

|  | **Cohen’s *d* [95% CI]** | | ***p*-value** | ***p*_FDR_** | ***n* (OCD)** | ***n* (HC)** |
| --- | --- | --- | --- | --- | --- | --- |
| Ventral | | 0.036 [-0.130, 0.202] | 0.673 | 0.841 | 314 | 253 |
| Pulvinar | | -0.100 [-0.268, 0.064] | 0.234 | 0.455 | 312 | 252 |
| Medial | | -0.014 [-0.179, 0.151] | 0.868 | 0.868 | 316 | 253 |
| Lateral | | 0.120 [-0.041, 0.290] | 0.145 | 0.455 | 316 | 252 |
| Anterior | | 0.094 [-0.073, 0.260] | 0.273 | 0.455 | 311 | 251 |
| Whole Thal | | -0.020 [-0.185, 0.145] | 0.811 | - | 317 | 253 |

Note: models were adjusted for age, sex, and intracranial volume; 95% CI = 95% Confidence Interval; FDR = False Discovery Rate; *n* = number of participants; OCD = Obsessive-Compulsive Disorder; HC = Healthy Controls; Thal = Thalamus.

**Table S22: Sex-by-diagnosis interaction in adult obsessive-compulsive patients and healthy controls**

|  | **Cohen’s *d* [95% CI]** | | ***p*-value** | ***p*_FDR_** | ***n* (OCD)** | ***n* (HC)** |
| --- | --- | --- | --- | --- | --- | --- |
| Ventral | | -0.055 [-0.112, 0.003] | 0.065 | 0.164 | 2200 | 2383 |
| Pulvinar | | -0.048 [-0.105, 0.010] | 0.108 | 0.18 | 2211 | 2381 |
| Medial | | 0.006 [-0.052, 0.064] | 0.846 | 0.846 | 2198 | 2390 |
| Lateral | | -0.020 [-0.078, 0.037] | 0.489 | 0.611 | 2201 | 2402 |
| Anterior | | -0.066 [-0.124, -0.008] | 0.026 | 0.132 | 2195 | 2400 |
| Whole Thal | | -0.042 [-0.099, 0.015] | 0.152 | - | 2265 | 2402 |

Note: models were adjusted for age, sex, and intracranial volume; 95% CI = 95% Confidence Interval; FDR = False Discovery Rate; *n* = number of participants; OCD = Obsessive-Compulsive Disorder; HC = Healthy Controls; Thal = Thalamus.

**Table S23: Age-by-diagnosis interaction in adult obsessive-compulsive patients and healthy controls**

|  | **Cohen’s *d* [95% CI]** | ***p*-value** | ***p*_FDR_** | ***n* (OCD)** | ***n* (HC)** |
| --- | --- | --- | --- | --- | --- |
| Ventral | -0.009 [-0.067, 0.049] | 0.763 | 0.954 | 2200 | 2383 |
| Pulvinar | 0.016 [-0.042, 0.074] | 0.589 | 0.954 | 2211 | 2381 |
| Medial | -0.002 [-0.06, 0.056] | 0.955 | 0.955 | 2198 | 2390 |
| Lateral | -0.032 [-0.09, 0.025] | 0.273 | 0.682 | 2201 | 2402 |
| Anterior | -0.042 [-0.1, 0.016] | 0.157 | 0.682 | 2195 | 2400 |
| Whole Thal | 0.002 [-0.056, 0.059] | 0.957 | - | 2265 | 2402 |

Note: models were adjusted for age, sex, and intracranial; 95% CI = 95% Confidence Interval; FDR = False Discovery Rate; *n* = number of participants; OCD = Obsessive-Compulsive Disorder; HC = Healthy Controls; Thal = Thalamus.

**Table S24: Age-squared-by-diagnosis interaction in adult obsessive-compulsive patients and healthy controls**

|  | **Cohen’s *d* [95% CI]** | | ***p*-value** | ***p*_FDR_** | ***n* (OCD)** | ***n* (HC)** |
| --- | --- | --- | --- | --- | --- | --- |
| Ventral | | -0.009 [-0.067, 0.049] | 0.763 | 0.954 | 2200 | 2383 |
| Pulvinar | | 0.016 [-0.042, 0.074] | 0.589 | 0.954 | 2211 | 2381 |
| Medial | | -0.002 [-0.06, 0.056] | 0.955 | 0.955 | 2198 | 2390 |
| Lateral | | -0.032 [-0.09, 0.025] | 0.273 | 0.682 | 2201 | 2402 |
| Anterior | | -0.042 [-0.100, 0.016] | 0.157 | 0.682 | 2195 | 2400 |
| Whole Thal | | 0.002 [-0.056, 0.059] | 0.957 | - | 2265 | 2402 |

Note: models were adjusted for age, sex, and intracranial volume; 95% CI = 95% Confidence Interval; FDR = False Discovery Rate; *n* = number of participants; OCD = Obsessive-Compulsive Disorder; HC = Healthy Controls; Thal = Thalamus

**Table S25: Age-by-sex-by-diagnosis interaction in adult obsessive-compulsive patients and healthy controls**

|  | | **Cohen’s *d* [95% CI]** | ***p*-value** | ***p*_FDR_** | ***n* (OCD)** | ***n* (HC)** |
| --- | --- | --- | --- | --- | --- | --- |
| Ventral | 0.043 [-0.015, 0.101] | | 0.148 | 0.164 | 2200 | 2383 |
| Pulvinar | 0.054 [-0.003, 0.112] | | 0.065 | 0.180 | 2211 | 2381 |
| Medial | 0.036 [-0.022, 0.094] | | 0.224 | 0.846 | 2198 | 2390 |
| Lateral | 0.010 [-0.048, 0.068] | | 0.743 | 0.611 | 2201 | 2402 |
| Anterior | 0.002 [-0.056, 0.06] | | 0.941 | 0.132 | 2195 | 2400 |
| Whole Thal | 0.042 [-0.016, 0.099] | | 0.154 | - | 2265 | 2402 |

Note: models were adjusted for age, sex, and intracranial volume; 95% CI = 95% Confidence Interval; FDR = False Discovery Rate; *n* = number of participants; OCD = Obsessive-Compulsive Disorder; HC = Healthy Controls; Thal = Thalamus

**Table S26: Multiple linear regression output of volumetric difference between adult-onset obsessive-compulsive patients and healthy controls**

|  | **Cohen's *d* [95% CI]** | ***p*-value** | ***p*_FDR_** | ***n* (Med)** | ***n* (Unmed)** |
| --- | --- | --- | --- | --- | --- |
| Ventral | -0.053 [-0.127, 0.022] | 0.1660 | 0.1660 | 981 | 2383 |
| Pulvinar | -0.068 [-0.142, 0.006] | 0.0720 | 0.0890 | 989 | 2381 |
| Medial | -0.097 [-0.172, -0.023] | 0.0100 | 0.0260 | 982 | 2390 |
| Lateral | -0.082 [-0.156, -0.008] | 0.0310 | 0.0510 | 986 | 2402 |
| Anterior | -0.140 [-0.214, -0.065] | 0.0002 | 0.0010 | 980 | 2400 |
| Whole Thal | -0.085 [-0.159, -0.012] | 0.0230 | - | 1000 | 2402 |

Note: models were adjusted for age, sex, and intracranial volume; 95% CI = 95% Confidence Interval; FDR = False Discovery Rate; *n* = number of participants; OCD = Obsessive-Compulsive Disorder; HC = Healthy Controls; Thal = Thalamus

**Table S27: Multiple linear regression output of volumetric difference between child-onset obsessive-compulsive patients healthy controls**

|  | **Cohen's *d* [95% CI]** | ***p*-value** | ***p*_FDR_** | ***n* (OCD)** | ***n* (HC)** |
| --- | --- | --- | --- | --- | --- |
| Ventral | -0.015 [-0.094, 0.063] | 0.700 | 0.700 | 835 | 2383 |
| Pulvinar | -0.038 [-0.117, 0.040] | 0.34 | 0.609 | 840 | 2381 |
| Medial | -0.028 [-0.107, 0.051] | 0.487 | 0.609 | 832 | 2390 |
| Lateral | -0.034 [-0.113, 0.045] | 0.400 | 0.609 | 833 | 2402 |
| Anterior | -0.074 [-0.153, 0.005] | 0.065 | 0.325 | 836 | 2400 |
| Whole Thal | -0.045 [-0.123, 0.034] | 0.264 | - | 850 | 2402 |

Note: models were adjusted for age, sex, and intracranial volume; 95% CI = 95% Confidence Interval; FDR = False Discovery Rate; *n* = number of participants; OCD = Obsessive-Compulsive Disorder; HC = Healthy Controls; Thal = Thalamus

**Table S28: Multiple linear regression output of volumetric difference between adult-onset patients and child-onset obsessive-compulsive patients**

|  | **Cohen's *d* [95% CI]** | ***p*-value** | ***p*_FDR_** | ***n* (AO)** | ***n* (CO)** |
| --- | --- | --- | --- | --- | --- |
| Ventral | -0.033 [-0.126, 0.059] | 0.481 | 0.566 | 981 | 835 |
| Pulvinar | -0.027 [-0.119, 0.065] | 0.566 | 0.566 | 989 | 840 |
| Medial | -0.075 [-0.168, 0.017] | 0.110 | 0.551 | 982 | 832 |
| Lateral | -0.041 [-0.133, 0.051] | 0.384 | 0.566 | 986 | 833 |
| Anterior | -0.053 [-0.145, 0.040] | 0.265 | 0.566 | 980 | 836 |
| Whole Thal | -0.041 [-0.132, 0.051] | 0.385 | - | 1000 | 850 |

Note: models were adjusted for age, sex, and intracranial volume; 95% CI = 95% Confidence Interval; FDR = False Discovery Rate; *n* = number of participants; AO = Adult-onset obsessive-compulsive disorder; CO = Child-onset obsessive-compulsive disorder; Thal = Thalamus.

**Table S29: Multiple linear regression output of the association between severity (Child version Yale-Brown Obsessive Compulsive Scale) and volume in pediatric obsessive-compulsive patients**

|  | | **Pearson's *r*** | ***t*** | **df** | ***p*-value** | ***p*_FDR_** |
| --- | --- | --- | --- | --- | --- | --- |
| Ventral | 0.091 | | 0.859 | 89 | 0.393 | 0.491 |
| Pulvinar | 0.106 | | 1.004 | 89 | 0.318 | 0.491 |
| Medial | -0.014 | | -0.128 | 88 | 0.898 | 0.898 |
| Lateral | 0.207 | | 1.985 | 88 | 0.050 | 0.251 |
| Anterior | 0.158 | | 1.503 | 88 | 0.136 | 0.341 |
| Whole Thal | 0.141 | | 1.347 | 90 | 0.181 | - |

Note: Partial correlation is adjusted for age, sex, and intracranial volume; 95% FDR = False Discovery Rate; df = degrees of freedom; Thal = Thalamus.

**Table S30: Multiple linear regression output of the association between severity ((Child version) Yale-Brown Obsessive Compulsive Scale) and volume in adolescent obsessive-compulsive patients.**

|  | | **Pearson's *r*** | ***t*** | **df** | ***p*-value** | ***p*_FDR_** |
| --- | --- | --- | --- | --- | --- | --- |
| Ventral | 0.014 | | 0.247 | 302 | 0.805 | 0.851 |
| Pulvinar | -0.033 | | -0.571 | 307 | 0.568 | 0.851 |
| Medial | -0.011 | | -0.192 | 307 | 0.848 | 0.851 |
| Lateral | 0.011 | | 0.189 | 303 | 0.851 | 0.851 |
| Anterior | 0.034 | | 0.601 | 305 | 0.548 | 0.851 |
| Whole Thal | 0.014 | | 0.249 | 308 | 0.803 | - |

Note: Partial correlation is adjusted for age, sex, and intracranial volume; 95% FDR = False Discovery Rate; df = degrees of freedom; Thal = Thalamus.

**Table S31: Multiple linear regression output of the association between severity (Yale-Brown Obsessive Compulsive Scale) and volume in adult obsessive-compulsive patients.**

|  | **Pearson's *r*** | ***t*** | **df** | ***p*-value** | ***p*_FDR_** |
| --- | --- | --- | --- | --- | --- |
| Ventral | -0.054 | -2.443 | 2072 | 0.015 | 0.018 |
| Pulvinar | -0.045 | -2.048 | 2078 | 0.041 | 0.041 |
| Medial | -0.071 | -3.239 | 2075 | 0.001 | 0.003 |
| Lateral | -0.062 | -2.832 | 2088 | 0.005 | 0.008 |
| Anterior | -0.073 | -3.354 | 2078 | 0.001 | 0.003 |
| Whole Thal | -0.066 | -3.049 | 2113 | 0.002 | - |

Note: Partial correlation is adjusted for age, sex, and intracranial volume; 95% FDR = False Discovery Rate; df = degrees of freedom; Thal = Thalamus.

**Table S32: Multiple linear regression output of the asymmetry index differences between pediatric obsessive-compulsive patients and healthy controls**

|  | **Cohen's *d* [95% CI]** | ***p*-value** | ***p*_FDR_** | ***n* (OCD)** | ***n* (HC)** |
| --- | --- | --- | --- | --- | --- |
| Ventral | 0.180 [-0.111, 0.471] | 0.233 | 0.338 | 94 | 88 |
| Pulvinar | -0.140 [-0.436, 0.147] | 0.338 | 0.338 | 94 | 88 |
| Medial | 0.160 [-0.133, 0.451] | 0.293 | 0.338 | 94 | 87 |
| Lateral | 0.260 [-0.032, 0.549] | 0.086 | 0.338 | 95 | 89 |
| Anterior | 0.160 [-0.130, 0.448] | 0.288 | 0.338 | 95 | 90 |
| Whole Thal | 0.075 [-0.212, 0.363] | 0.613 | - | 96 | 90 |

Note: models were adjusted for age, sex, and intracranial volume; 95% CI = 95% Confidence Interval; FDR = False Discovery Rate; *n* = number of participants; OCD = Obsessive-Compulsive Disorder; HC = Healthy Controls; Thal = Thalamus.

**Table S33: Multiple linear regression output of the asymmetry index differences between adolescent obsessive-compulsive patients and healthy controls**

|  | **Cohen's *d* [95% CI]** | ***p*-value** | ***p*_FDR_** | ***n* (OCD)** | ***n* (HC)** |
| --- | --- | --- | --- | --- | --- |
| Ventral | 0.104 [-0.062, 0.270] | 0.223 | 0.557 | 311 | 251 |
| Pulvinar | 0.134 [-0.032, 0.300] | 0.115 | 0.557 | 316 | 252 |
| Medial | 0.000 [-0.166, 0.165] | 0.998 | 0.998 | 316 | 253 |
| Lateral | -0.037 [-0.203, 0.129] | 0.664 | 0.984 | 312 | 252 |
| Anterior | -0.023 [-0.189, 0.143] | 0.787 | 0.984 | 314 | 253 |
| Whole Thal | -0.017 [-0.182, 0.148] | 0.837 | - | 317 | 254 |

Note: models were adjusted for age, sex, and intracranial volume; 95% CI = 95% Confidence Interval; FDR = False Discovery Rate; *n* = number of participants; OCD = Obsessive-Compulsive Disorder; HC = Healthy Controls; Thal = Thalamus

**Table S34: Multiple linear regression output of the asymmetry index differences between adult obsessive-compulsive patients and healthy controls**

|  | **Cohen's *d* [95% CI]** | ***p*-value** | ***p*_FDR_** | ***n* (OCD)** | ***n* (HC)** |
| --- | --- | --- | --- | --- | --- |
| Ventral | 0.025 [-0.033, 0.083] | 0.398 | 0.498 | 2200 | 2383 |
| Pulvinar | 0.046 [-0.012, 0.104] | 0.120 | 0.200 | 2211 | 2381 |
| Medial | 0.05 [-0.008, 0.108] | 0.089 | 0.200 | 2198 | 2390 |
| Lateral | 0.014 [-0.043, 0.072] | 0.627 | 0.627 | 2201 | 2402 |
| Anterior | -0.058 [-0.116, 0.000] | 0.051 | 0.200 | 2195 | 2400 |
| Whole Thal | 0.041 [-0.017, 0.098] | 0.165 | - | 2236 | 2402 |

Note: models were adjusted for age, sex, and intracranial volume; 95% CI = 95% Confidence Interval; FDR = False Discovery Rate; *n* = number of participants; OCD = Obsessive-Compulsive Disorder; HC = Healthy Controls; Thal = Thalamus
